# Tight Fit of the SIR Dynamic Epidemic Model to Daily Cases of COVID-19 Reported During the 2021-2022 Omicron Surge in New York City: A Novel Approach

**DOI:** 10.1101/2023.03.13.23287177

**Authors:** Jeffrey E. Harris

**Author notes:** See Author Declarations at the end of the manuscript for additional details.

## Abstract

We describe a novel approach to recovering the underlying parameters of the SIR dynamic epidemic model from observed data on case incidence. We formulate a discrete-time approximation to the original continuous-time model and search for the parameter vector that minimizes the standard least squares criterion function. We show that the gradient vector and matrix of second-order derivatives of the criterion function with respect to the parameters adhere to their own systems of difference equations and thus can be exactly calculated iteratively. Applying our new approach, we estimated a four-parameter SIR model from data on daily reported cases of COVID-19 during the SARS-CoV-2 Omicron/BA.1 surge of December 2021 - March 2022 in New York City. The estimated SIR model showed a tight fit to the observed data, but less so when we excluded residual cases attributable to the Delta variant during the initial upswing of the wave in December. Our analyses of both the real-world COVID-19 data and simulated case incidence data revealed an important problem of weak parameter identification. While our methods permitted separate estimation of the infection transmission parameter and the infection persistence parameter, only a linear combination of these two key parameters could be estimated with precision. The SIR model appears to be an adequate reduced-form description of the Omicron surge, but it is not necessarily the correct structural model. Prior information above and beyond case incidence data may be required to sharply identify the parameters and thus distinguish between alternative epidemic models.

## 1. Introduction

Nowadays our sophisticated graphic software can draw attractive plots showing how many people have fallen victim to a highly contagious disease over the course of days, weeks, or months. But our graphs alone don’t teach us how to reliably determine the underlying risk of transmission from an infected to a susceptible person, or the amount of time that an infected individual remains contagious to others, or what proportion of the population was already infected at the critical point in time when the epidemic wave took off, or how many people remain at risk of infection.

We’ve just described what mathematicians call the inversion problem [1-4]: how to work backwards from limited data on incident cases or deaths to recover the key parameters underlying our dynamic epidemic models. The problem was born nearly a century ago when Kermack and McKendrick (KM) fit a curve derived from their now-classic model to datapoints of weekly deaths from a plague outbreak on the Isle of Bombay [5]. Since then, scores of investigators have searched for a robust, workable method of estimating the parameters of what has famously come to be known as the SIR (Susceptible-Infected-Removed) model, and the race to find a solution has accelerated with the arrival of the COVID-19 epidemic.

What has made the inversion problem so difficult is that, with some possible exceptions [6-10], the SIR model of coupled differential equations does not admit a closed-form mathematical solution that can be readily used to test the model’s predictions against the observed data. That major stumbling block has left us with a motley collection of second-best alternatives.

One idea has been to back out the parameters from the certain salient characteristics of the observed epidemic curve, such as the initial exponential rate of increase of cases [11, 12], the time to reach the peak incidence [13], the rate of decline after the peak [14], and the proportion of the population that is ultimately infected [15-17]. This approach may give us point estimates of the key parameters, but it does not provide any uncertainty ranges around the estimates.

Another idea is to pare down the set of parameters to be identified by making judicious use of prior information on some parameters [18-21], in some cases derived from previous waves of a multi-wave epidemic [22]. Perhaps the most traveled road to a solution has been the use of various parameter search algorithms [18, 23-29] which, when it comes down to it, offer only a marginal improvement over brute force search [30]. Bayesian estimation may be better able to integrate prior information into our search procedures [31-36], but its computational burden is usually even greater. Last but not least, we can resort to trial and error combined with visual inspection [37].

The present study, we suggest, offers an easily workable solution to the inversion problem. Rather than seeking a closed-form, analytical solution to KM’s system of differential equations, we pursue an alternate strategy. First, following the lead of other investigators [35, 38-40], we develop a discrete-time version of their classic, continuous-time SIR model. This step allows us to write their dynamic system in terms of difference equations rather than differential equations. Second, similarly following in others’ footsteps [26, 41-44], we define a least squares objective function to test our SIR model’s predictions against the observed data.

Third, in what appears to be an innovation, we show that both the gradient vector and Hessian matrix of second-order derivatives of our objective function with respect to the parameters follow their own systems of difference equations. As a result, both the gradient and Hessian can be rapidly and exactly computed by straightforward iteration, an approach that is computationally superior to numerical approximation [45].

Fourth, once we have calculated the gradient and Hessian, we can use the well-known Newton-Raphson algorithm [46] to find the global optimum. Fifth, relying on the minimum least squares criterion, we can then calculate the variance-covariance matrix of the parameters and thus determine their confidence intervals [47]. Sixth, our approach permits us to readily determine what parameters are in fact identified when we have only time-series data on new cases.

We apply our strategy to the estimation of a four-parameter SIR model to study COVID-19 incidence over a 99-day interval from the December 4, 2021, through March 12, 2022, during the Omicron/BA.1 wave in New York City.

## 2. Statistical Methods

### 2.1. Discrete-Time SIR Model

Following the lead of other investigators [4, 35, 38-40], we adopt a discrete-time approach. We mark off the time axis in equally spaced intervals *t* = 0, 1, …, *T*, where the duration of each interval is sufficiently small as to adequately approximate the classical, continuous-time version [5, 48-51]. At any time *t*, individuals within this closed population can be in one of three mutually exclusive states: *susceptible* (*S*), *infected* (*I*), or *removed* (*R*). The latter state, which includes both recovered living individuals and decedents, is assumed to be absorbing. To minimize possible complications arising from the non-identifiability of multiple parameters [52-55], we assume a fixed, demographically closed population of size *N*.

Let *S*_*t*_, *I*_*t*_, and *R*_*t*_ denote the respective numbers of individuals in each of the three states at time *t*. The dynamic path of the epidemic is governed by the following deterministic, coupled difference equations:

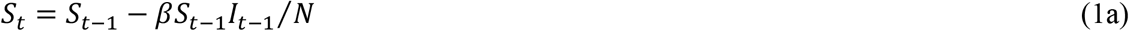

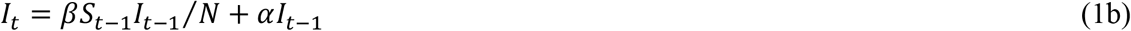

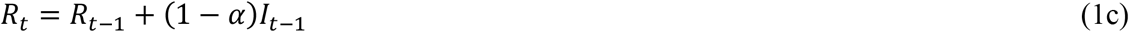

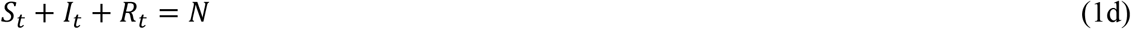

Equations (1a) through (1d) represent the well-known forward Euler approximation to the underlying continuous-time SIR model of coupled differential equations [56]. Apart from the population size *N*, this dynamic system has two parameters: *β* and *α*. In equations (1a) and (1b), *β* > 0 is an infection transmission parameter. In equations (1b) and (1c), 1 > *α* > 0 is an infection persistence parameter. It gauges the proportion of infected individuals at each time *t* who remain in the infected state. The quantity (1 − *α*) in equation (1c), which in some treatments is represented by the parameter *γ*, corresponds to the proportion who transition to the removed state. The multiplicative term *βS*_*t*−1_*I*_*t*−1_/*N* in equations (1a) and (1b) reflects the law of mass action [57], whereby susceptible individuals become infected in proportion to their frequency of contact with currently infected individuals. All individuals within the population are assumed to mix homogeneously, with no subgroup of individuals mixing preferentially with any other subgroup.

The final equation (1d) reflects the constant size *N* of the population and is consistent with equations (1a) through (1c). Strictly speaking, one should adjust the size *N* of the mixing population in equations (1a) and (1b) to take account of removals by death. Unless the overall death rate is substantial, this adjustment is usually ignored in model implementations.

We complete our discrete-time model with the specification of initial conditions at *t* = 0:

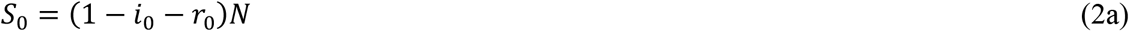

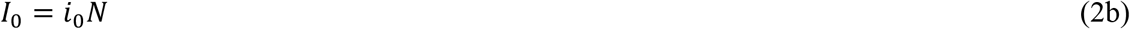

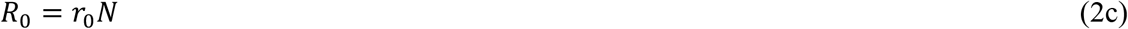

The additional parameters *i*_0_ and *r*_0_ represent the respective proportions of the entire population of size *N* that are initially infected and removed.

In the dynamic system (1) and (2), the mean duration of infection is time-invariant and equal to 1/(1 − *α*). The system results in an epidemic wave when (1 − *i*_0_ − *r*_0_) *β*/(1 − *α*) > 1, a well-known result known as the epidemic threshold theorem [24, 50, 58, 59]. Assuming that 1 − *i*_0_ − *r* _0_ ≈ 1, most authors write this epidemic threshold condition as ℛ_0_ > 1, where ℛ_0_ = *β*/(1 − *α*) is defined as the basic reproduction number [24, 51, 60, 61].

### 2.2. Parameter Estimation: Least Squares Minimization Criterion

We do not have direct observations on the underlying *state variables S*_*t*_, *I*_*t*_, and *R*_*t*_. If we had such data, our inversion problem would border on trivial [39, 62, 63]. Instead, we observe only the reported counts of new infections at various intervals. In this exposition, we assume that new infection counts *y*_*t*_ are observed at each discrete time *t*. The more general case where such counts are observed less frequently is discussed below. The counts *y*_*t*_ represent observations on the *output variables X*, *t* = 1, …, *T*, which from (1) correspond to:

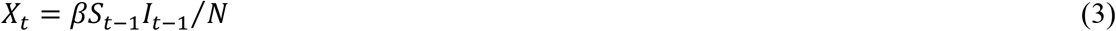

Given the definition of the output variable *X*_*t*_ in (3), our dynamic system (1) can be redefined as:

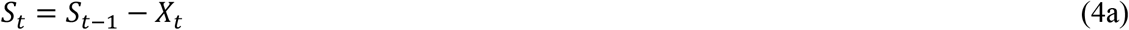

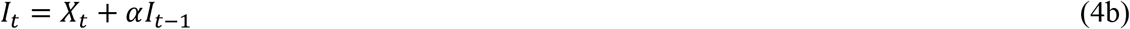

So long as the state variables adhere to the condition that that *S*_*t*_ + *I*_*t*_ + *R*_*t*_ = *N*, an explicit difference equation for *R*_*t*_ is unnecessary.

We can now characterize our parameter estimation problem. Given observations *y*_*t*_ on the output variables *X*_*t*_, we want to estimate the unknown parameters *β, α, i*_0_, *r*_0_ and *N*. Our stumbling block is that we cannot express *X*_*t*_ as a closed-form function of these parameters. We know only that the output variables *X*_*t*_ adhere to the dynamic system defined by (2), (3) and (4), which in turn depends on these unknown parameters.

Let ***y*** = (*y*_1_, …, *y*_*T*_)′ and ***X*** = (*X*_1_, …, *X*_*T*_)′, respectively, denote column vectors of the observed incidence data and the corresponding output variables at each time *t*, where we use boldface symbols denote vectors or matrices. Let **Θ** = (*β, α, i*_0_, *r*_0_, *N*) denote the row vector of the unknown parameters. Let ***X***(**Θ**) represent the functional dependence of the output variables on these parameters. To estimate **Θ** from the data ***y***, we introduce the least squares minimization criterion *V*(**Θ**) = ***y*** − ***X***(**Θ**)′(***y*** − ***X***(**Θ**)), which can be written in summation notation as:

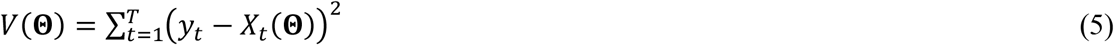

This criterion has been widely used in attempts to fit the SIR model to incidence data [26, 41-44]. It is well known that minimizing *V* is equivalent to computing the maximum likelihood estimate of **Θ** under the assumption that *y*_*t*_ = *X*_*t*_(**Θ**) + *ε*_*t*_, where the disturbances ε_*t*_ are independently normally distributed *N*(0, *σ*^2^) with homoscedastic variance *σ*^2^.

### 2.3. Non-Identifiability of the Five-Parameter Vector Θ

While the model delineated in equations (1) through (4) is standard in the literature [4, 35, 38-40], it turns out that not all five parameters of the vector **Θ** = (*β, α, i*_0_, *r*_0_, *N*) can be identified from the data ***y*** on case incidence alone. This conclusion is supported by the following proposition, which, along with all other propositions, is proved in Appendix A.

#### Proposition 1.

Let Ω ⊆ ℝ^5^ be the subspace of admissible values of the five-dimensional parameter vector **Θ**. Let ***X***(**Θ**) be the resulting *T* × 1 column vector of output variables defined by the model of equations (1) through (4). Then there exists a mapping *ϕ*: Ω → ℝ^4^ with the property that ***X***(**Θ**) = ***X***(**Θ**′) for all vectors **Θ, Θ**′ ∈ Ω satisfying *ϕ*(**Θ**) = *ϕ*(**Θ**′).

Even if we had perfectly accurate data ***y*** on case incidence, we could no better than to measure ***X*** without error. Proposition 1, however, establishes that ***X***(**Θ**) is not a one-to-one mapping from parameters **Θ** to output variables ***X***. Moreover, for any given set of observations ***y*** on case incidence, equation (5) teaches us that the dependence of *V* on **Θ** runs solely through ***X***(**Θ**). Accordingly, *V*(**Θ**) is likewise not a one-to-one mapping from parameters **Θ** to our objective function *V*.

Appended to the proof of Proposition 1 are some corollary results indicating how additional prior information can be used to identify the model parameters. For example, if we had sharp prior information on the initial proportion of recovered individuals *r*_0_, or the initial population size *N*, or the basic reproduction number ℛ_0_, then we could identify all the parameters in **Θ**. In what follows, we impose the prior constraint that *r*_0_ = 0. To simplify our notation, we drop *r*_0_ from the parameter vector and instead write **Θ** = (*β, α, i*_0_, *N*) as a four-parameter vector, denoting by Ω ⊆ ℝ^4^ the four-dimensional space of admissible values of **Θ**.

### 2.4. Gradient and Hessian of the Least Squares Criterion V

Let the operator ***D*** denote the 4 × 1 gradient of partial derivatives with respect to the elements of **Θ**, and let the operator ***D***^**2**^ denote the corresponding 4 × 4 Hessian matrix of second order partial derivatives. The following two propositions embody the main methodological innovation of this paper.

#### Proposition 2.

The gradient of the least squares criterion *V* with respect to the parameter vector **Θ** = (*β, α, i*_0_, *N*) is:

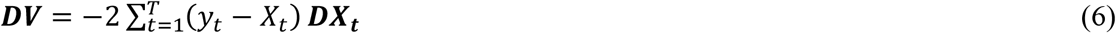

where each column vector 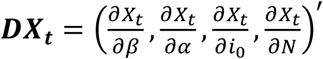 represents the corresponding gradient of partial derivatives of the output variable *X*_*t*_ at time *t*. The basic equations of our dynamic system (1), in combination with the initial conditions (2), can be used to generate complete, computable difference equations for ***DX***_***t***_ for all *t*, and thus for ***DV***.

#### Proposition 3.

Let ***DX*** denote the *T* × 4 matrix whose *t*-th row is the vector 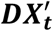, as defined in Proposition 2. The Hessian matrix of the least squares criterion *V* with respect to the parameter vector **Θ** = (*β*, *α*, *i* _0_, *N*) is:

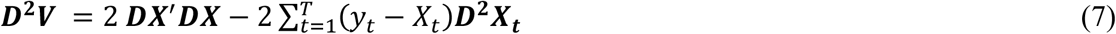

The basic equations of our dynamic system (1), in combination with the initial conditions (2), can similarly be used to generate complete, computable difference equations for ***D***^**2**^***X***_***t***_ for all *t*, and thus ***D***^**2**^***V***.

As part of the proofs of these two propositions in Appendix A, we show the details of the difference equations involved in the exact computation of ***DX***_***t***_ and ***D***^**2**^***X***_***t***_ for all *t*. We also address the case where observations *y*_*t*_ are not available for all times *t*.

### 2.5. Newton-Raphson Algorithm for Parameter Estimation

While the objective function *V*(**Θ**) has no closed-form expression in terms of **Θ**, we can still rely on our exact computations of the gradient ***DV*** and Hessian ***D***^**2**^***V*** to search for a local interior minimum within the four-dimensional subspace Ω of admissible values of **Θ** via the Newton-Raphson algorithm [46].

The Newton-Raphson algorithm is iterative. Denote our choice of initial parameter vector as **Θ**^(0)^. Let (*k*) index successive iterations. The current value of the parameter vector **Θ**^(*k*)^ is repeatedly mapped into an updated value **Θ**^(*k*+1)^ according to the well-known rule:

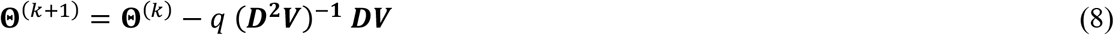

Here, the gradient ***DV***, as defined in (6), and the Hessian ***D***^**2**^***V***, as defined in (7), are both computed at the current value of the parameter vector **Θ**^(*k*)^, while the step size 0 < *q* ≤ 1 is under control of the programmer.

This iterative approach would work flawlessly if the objective function *V* were globally convex with an interior minimum. There is good reason, however, to suspect that *V* may instead have multiple local optima. While the classical SIR model predicts a single-peaked wave of incident cases ***X*** so long as ℛ_0_ > 1, the observed data ***y*** often display multiple peaks over time. (A good example is the two-peak plot of the 2001 Dengue fever outbreak in Havana [59].) In that case, the search algorithm embodied in (8) may easily end up at a local rather than a global minimum of *V*. To address this possibility, we need to run the Newton-Raphson search routine from various initial parameter vectors **Θ**^(0)^.

What’s more, there may be regions of the four-parameter space where the Hessian ***D***^**2**^***V*** is not positive definite because the second term in equation (7) (that is, 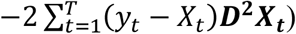) is not necessarily a positive definite matrix. In that case, the parameter updating rule (8) may not result in a decrease in the objective function *V* as the algorithm veers away from the optimum. To address this possibility, we can back up to **Θ**^(*k*)^ and reduce the step size *q*.

### 2.6. Alternative EM-Type Procedure for Parameter Estimation

It turns out that there is an alternative iterative procedure for parameter estimation analogous to the so-called EM algorithm [64]. Rather than searching through the four-dimensional space of admissible values of **Θ** = (*β, α, i*_0_, *N*), we can separate the estimation of the population-size parameter *N* and from our search over the remaining three-dimensional space of the remaining identifiable parameters (*β, α, i*_0_). The procedure is motivated by the following two propositions.

#### Proposition 4.

Each element *X*_*t*_(**Θ**) of the output vector ***X***(**Θ**) is a linear function of the population size parameter *N*. That is, *X*_*t*_ can be written in the form *φ*_*t*_(***θ***)*N*, where *φ*_*t*_(***θ***) is a function of the remaining identifiable parameters ***θ*** = (*β, α, i*_0_).

This result does not require that we derive a closed-form expression for the function *φ*_*t*_(***θ***). The fact that each output variable *X*_*t*_ is proportional to *N* implies that the optimization criterion *V* defined in (5) is a quadratic polynomial in *N*. That conclusion in turn motivates the following additional proposition.

#### Proposition 5.

Let **Θ**^(*n*)^ = (***θ***^(*n*)^, *N*^(*n*)^) denote parameter estimates at iteration *n* of an iterative estimation algorithm. Let ***X***^(***n***)^ = ***X*(Θ**^(*n*)^) denote the corresponding output variable vector derived from the SIR model (2), (3) and (4) based upon these parameter estimates. Define

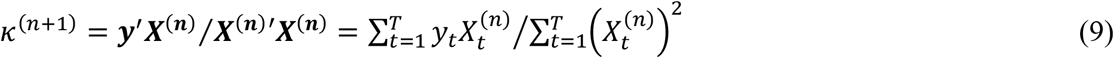

as the regression coefficient of ***y*** on ***X***^(***n***)^. Then conditional upon **Θ**^(*n*)^, the updated population-size parameter that minimizes the least squares criterion *V* is *N*^(*n*+1)^ = *κ*^(*n*+1)^*N*^(*n*)^.

Proposition 5 describes the E (or expectation) step for updating the estimate of the population-size parameter *N*^(*n*+1)^ in the iterative algorithm. The minimization (or M) step consists of determining the parameter vector ***θ***^(*n*+1)^ that minimizes the least squares criterion *V* conditional upon the updated value of *N*^(*n*+1)^. The latter step can be similarly performed via the Newton-Raphson algorithm, as described above, but where the parameter search is now confined to the three-dimensional space of ***θ*** = (*β, α, i*_0_).

### 2.7. Confidence Intervals

#### Proposition 6.

Let 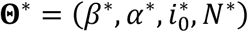 and denote the estimated parameter values that minimize the objective function *V*(**Θ**) under the identifying restriction *r*_0_ = 0, and let *V*\* = *V*(**Θ**\*) denote the corresponding minimized value of the least squares criterion *V*. Then the variance-covariance matrix of the estimated parameters **Θ**\* is ***C***\* = (*s*\*)^2^(***DX***\*′***DX***\*)^−1^, where (*s*\*)^2^ = *V*\*/*T*, and where ***DX***\*, as defined in Proposition 3, is also evaluated at the optimum **Θ**\*.

This result allows us to estimate confidence intervals for our parameter estimates 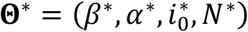 given the asymptotic normality of the least squares estimators derived here. Thus, the standard errors are the square roots of the diagonal elements of ***C***\*, while the symmetric 95 percent confidence intervals can be evaluated as ±1.96 standard errors about the estimates. We can then use the Delta method [65] to compute the corresponding standard errors and confidence intervals around nonlinear functions of the parameters, such as the basic reproduction number ℛ_0_ = *β*/(1 − *α*) and the mean duration of infection 1/(1 − *α*).

### 2.8. Only a Linear Combination of the Parameters β and α is Practically Identifiable

We have imposed the sharp prior restriction that *r*_0_ = 0 to identify the remaining four parameters of our SIR model. If there exists a local interior minimum 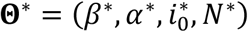 of our least squares objective function *V*(**Θ**), we know that the gradient ***DV***(**Θ**\*) is the zero vector and the Hessian ***D***^**2**^***V***(**Θ**\*) is a non-negative definite matrix. When **Θ**\* is a strict local minimum, ***D***^**2**^***V***(**Θ**\*) will be strictly positive definite.

Even if our objective function *V* is indeed *strictly* locally convex at **Θ**\* and thus ***D***^**2**^***V***(**Θ**\*) is strictly positive definite, it turns out that the Hessian ***D***^**2**^***V***(**Θ**\*) is *nearly* positive semidefinite. That is, *V*(**Θ**) is *nearly* flat along a ray passing through **Θ**\* where the parameters *β* and *α* are allowed to vary and the remaining parameters 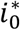 and *N*\* remain fixed. In effect, the three-dimensional plot of *V* projected onto the (*β, α*) plane contains a ravine where only a linear combination of *β* and *α* is *practically identified* [66].

The following proposition, which formalizes the idea, will guide our empirical implementation below.

#### Proposition 7.

Let **Θ**\* be a local interior minimum of *V*(**Θ**). Define a single-valued mapping from the real line into the four-dimensional subspace Ω of admissible values of **Θ** as follows: 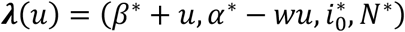, where *u* is its real-valued argument and *w* > 0 is a positive real parameter. As the parameter *w* varies, this mapping characterizes a family of rays in four-dimensional space passing through the point **Θ**\* at *u* = 0. Then there exists a ray, corresponding to a specific value of *w*, along which the second-order directional derivative 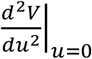 is nearly zero, that is, along which *V*(**Θ**) is *nearly* equal to its local minimum value *V*(**Θ**\*).

### 2.9. Consistency Checks with Data Simulated from Known Parameters

Apart from our study of COVID-19 case incidence in New York City, to be described below, we applied our estimation procedure to a simulated database generated from known parameters. Specifically, we computed the output variables *X*_*t*_ from the model of equations (2), (3) and (4) over *T* = 100 time periods with assumed known parameters **Θ** = (*β, α, i*_*o*_, *N*) = (0.6, 0.7, 0.04, 10^5^). We then repeatedly simulated the observed case incidence as 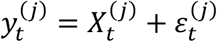 for each of *t* = 1, …, 100 time periods and *j* = 1, …, 100 epidemics, where the independent errors 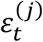 were randomly drawn from a Gaussian *N*(0, *σ*^2^) distribution with zero mean and standard deviation *σ* = 100. For each epidemic *j*, we then employed our 4-dimensional Newton-Raphson algorithm to estimate the parameters 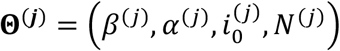. In addition, for each epidemic *jj*, we computed the 3-dimensional estimates of 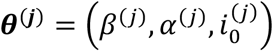 conditional upon the population size *N* known to equal its assumed value of 10^5^. Finally, for each of these exercises, we compared the dispersion of the resulting estimates **Θ**^(***j***)^ around their assumed values **Θ**.

### 2.10. Computational Benchmarks

We employed two versions of our procedure to estimate the parameters of our SIR models: the 4-parameter search Newton-Raphson search method embodied in Propositions 2 and 3; and the EM-type procedure embodied in Propositions 4 and 5. We compared the computational times attained through these two approaches with a brute-force, grid search over the 4-dimensional space of allowable values of **Θ**. All programs were written in Mata [67], a matrix programming language embedded within the Stata programming language [68]. Calculations were carried out on a MacBook Pro with a 2.3 GHz 8-Core Intel Core i9 processor.

## 3. Data

### 3.1. Omicron Wave, December 2021 – March 2022, New York City

We studied the reported daily incidence of COVID-19 during the SARS-CoV-2 Omicron/BA.1 wave of December 2021 – March 2022 in New York City, NY, United States, a city of population 8.5 million. Our data consisted of daily counts of cases reported by the New York City department of health [69], where the date of report was intended to be the date when a positive test was performed or when the diagnosis of COVID-19 was otherwise made.

Our data showed systematic variation in case counts by day of the week, with many fewer cases diagnosed over the weekends. To account for these fluctuations, and to accommodate delays between symptom onset and testing, we converted the raw case counts *c*_*t*_ into centered 7-day moving averages, that is, 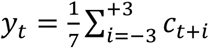, where *t* indexes the date of report. Figure 1 shows the raw counts of daily reported cases *c*_*t*_ (connected gray datapoints) as well as the daily case counts *y*_*t*_ adjusted for the day of the week (red datapoints). We relied upon these adjusted daily counts *y*_*t*_ to estimate our SIR model parameters **Θ** = (*β, α, i*_0_, *N*).

**Figure 1.**
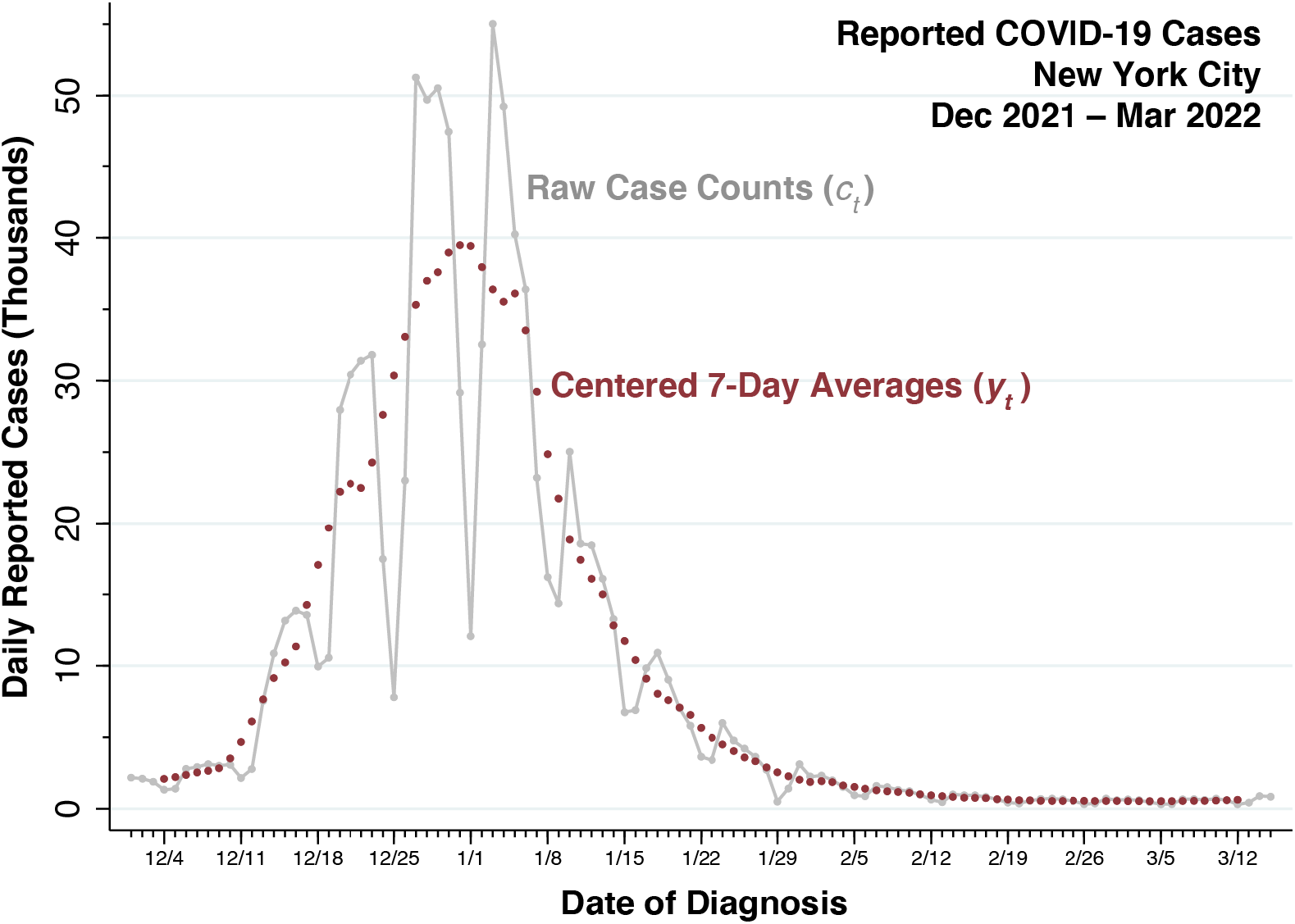
Daily Reported Cases of COVID-19, 12/1/2021 – 3/15/2022, in New York City. The connected gray datapoints show the raw case counts (*c*_*t*_). The red datapoints, covering 12/4/2021 – 3/12/2022, show the centered 7-day moving averages (*y*_*t*_).

The Omicron BA.1 variant of the SARS-CoV-2 virus was far-and-away the most important contributor to the massive surge of reported infections observed in Figure 1. Still, the initial phase of the surge overlapped the tail end of the prior Delta wave, while the terminal phase saw the gradual emergence of the Omicron BA.2 variant. Figure 2 plots the estimated daily proportions of the three variants in New York City during our study period. As an alternative analysis, we re-estimated our SIR model parameters **Θ** = (*β, α, i*_0_, *N*) from the data 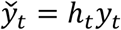, where *h*_*t*_ represents the estimated proportion of BA.1 infections on date *t*, as derived from the interpolated Omicron BA.1 curve in Figure 2, and where *y*_*t*_ are the moving average counts of all cases displayed in Figure 1.

**Figure 2.**
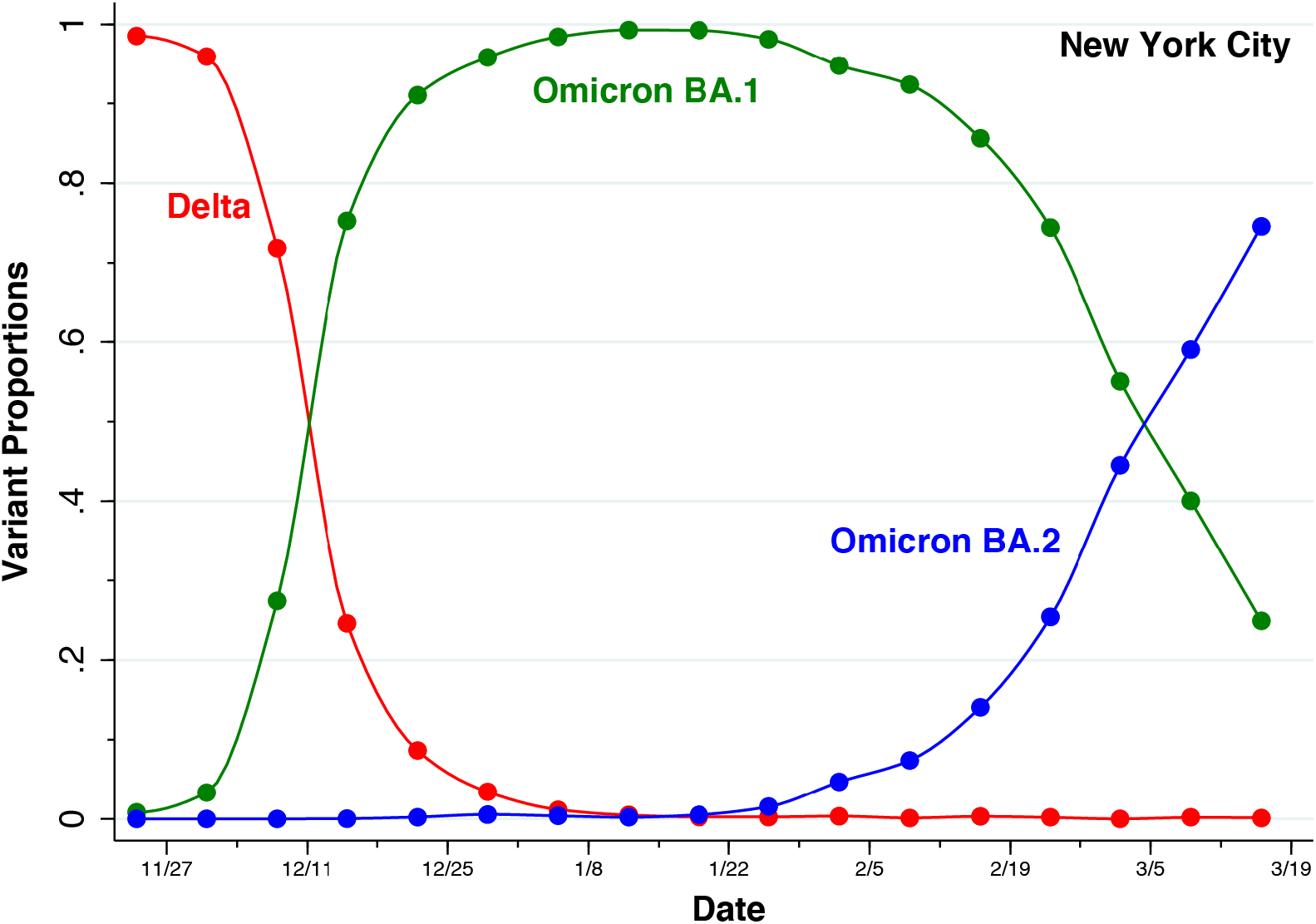
Estimates of the Proportions of the Delta, Omicron BA.1, and Omicron BA.2 Variants in New York City During 11/24/2021 – 3/16/2022. Weekly averages (solid datapoints) were derived from a compilation maintained by the New York City health department [70]. The Stata *pchipolate* interpolation routine [71] was then employed to estimate the intervening days (connecting curves). The Omicron BA.2 category included strains identified as BA.2, BA.2.12.1, and BA.2.75. Not shown are the proportions of all other strains, which represented no more than 0.7 percent of total samples in any one week.

## 4. Results

### 4.1. Omicron Wave, December 2021 – March 2022, New York City

Figure 3 shows the predicted values of the output variable *X*_*t*_ as connected curves superimposed on the observed datapoints *y*_*t*_ for all reported cases (shown in blue) and 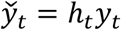 for Omicron BA.1 only cases (shown in orange). For all reported cases, the SIR model showed a reasonably tight fit to the data. For Omicron BA.1 only cases, however, the fitted SIR model overpredicted the initial number of cases during the first 10 days of observation (from 12/4 through 12/13/2021). Comparing the two fitted curves, we see that our dropping the Delta variant cases during December 2021 resulted in an attenuation of the initial upswing of epidemic wave. By contrast, elimination of the Omicron BA.2 cases resulted in only a small absolute decrease in the tail of the wave during February and early March of 2022.

**Figure 3.**
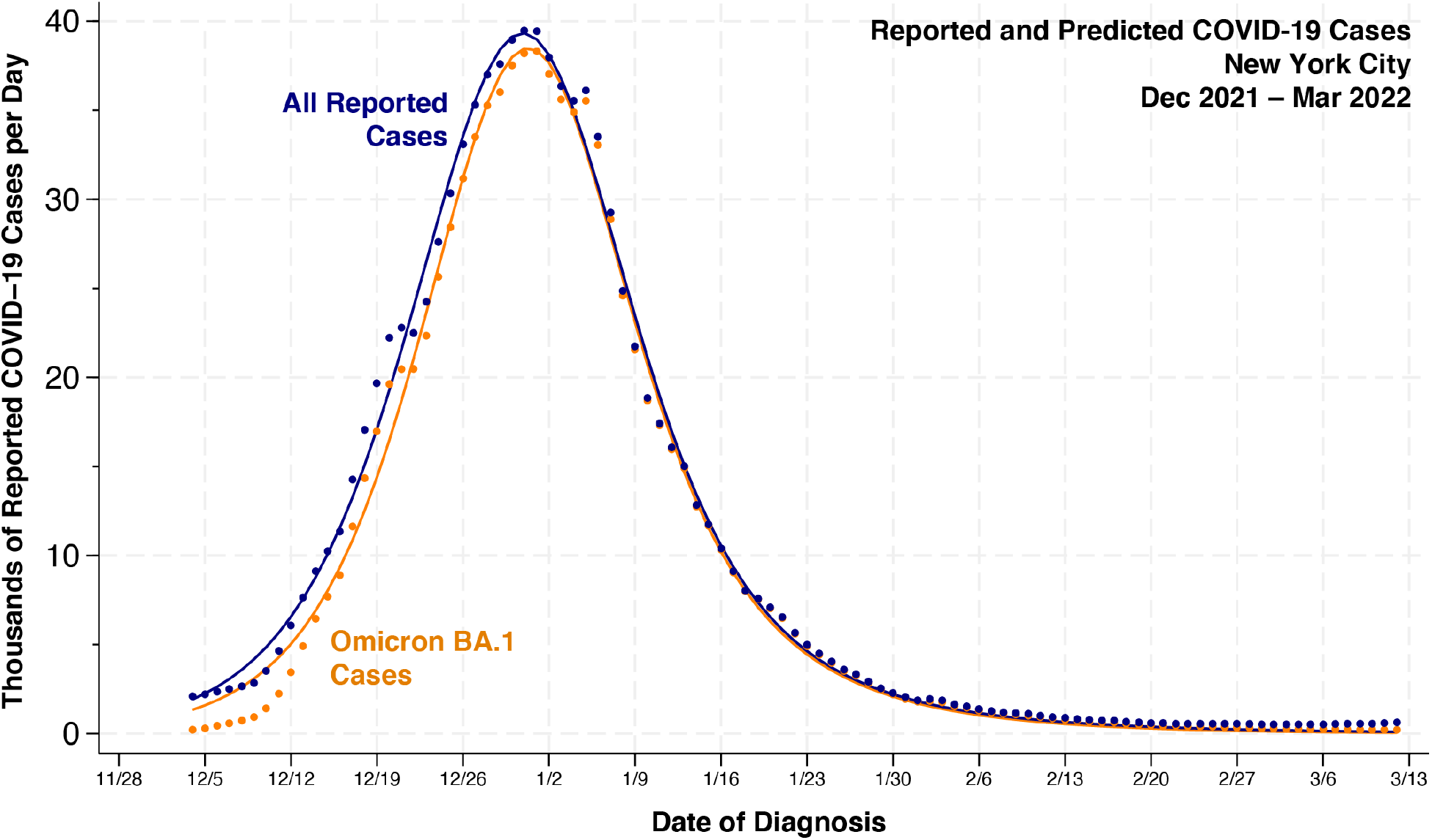
Daily Reported and Predicted Cases of COVID-19, 12/1/2021 – 3/15/2022, in New York City. Estimates Based Upon All Variants (Blue) and Omicron BA.1 Variant Only (Orange). Blue datapoints represent all reported cases *y*_*t*_, while orange datapoints represent Omicron BA.1 cases 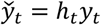. The respective blue and orange curves connect the predicted values of the output variable *X*_*t*_. The displayed estimates are based upon the 4-dimensional Newton-Raphson algorithm described in Propositions 2 and 3. The estimates based upon the alternative EM procedure, described in Propositions 4 and 5, were identical.

Table 1 summarizes the resulting parameter estimates. The 95% confidence intervals based upon the 4-dimensional Newton-Raphson approach, shown in parentheses below each point estimate, accounted for the uncertainty in all components of the parameter vector **Θ** = (*β, α, i*_0_, *N*). When we utilized the alternative EM-type algorithm, the final M-step estimated the three parameters (*β, α, i*_0_) *conditional* upon the value of *N* estimated from the prior E-step. The corresponding conditional confidence intervals, shown in square brackets, were somewhat tighter.

**Table 1.** Parameter Estimates of an SIR Model of COVID-19 Incidence New York City, December 2021 – March 2022. Data on All Cases Versus Omicron BA.1 Only ^a, b, c^.

|  |  |  |
| --- | --- | --- |
| **Parameter** | **All Reported Cases** | **Omicron BA.1 Only** |
| $\beta$ | 0.233<br>(0.200, 0.266)<br>[0.216, 0.250] | 0.283<br>(0.189, 0.377)<br>[0.266, 0.299] |
| $\alpha$ | 0.941<br>(0.912, 0.969)<br>[0.928, 0.953] | 0.903<br>(0.815, 0.991)<br>[0.890, 0.916] |
| $i_0 \times 10^{-3}$ | 8.23<br>(6.17, 10.28)<br>[6.95, 9.50] | 4.85<br>(2.20, 7.50)<br>[4.15, 5.56] |
| $N \times 10^6$ | 1.013<br>(0.980, 1.046)<br>— | 0.984<br>(0.868, 1.100)<br>— |
| $\mathcal{R}_0$ | 3.92<br>(2.58, 5.27)<br>[3.35, 4.50] | 2.92<br>(1.23, 4.61)<br>[2.70, 3.14] |
| $1/(1 - \alpha)$ | 16.8<br>(8.7, 25.0)<br>[13.2, 20.5] | 10.3<br>(0.9, 19.7)<br>[8.9, 11.7] |
| $V$ | 76.7686 | 87.3200 |
| $AIC$ | −15.1780 | −2.4284 |
| a. The 4-dimensional Newton-Raphson approach and the alternative EM-algorithm approach gave the same point estimates of the parameters $\Theta = (\beta, \alpha, i_0, N)$.<br>b. Symmetric 95% confidence intervals based upon the 4-dimensional Newton-Raphson algorithm are shown in parentheses below each estimate. The corresponding confidence intervals conditional upon the population size parameter $N$ are shown in square brackets. Confidence intervals for the estimated basic reproduction number $\mathcal{R}_0 = \beta/(1 - \alpha)$ and the estimated mean duration of infectivity $1/(1 - \alpha)$ were based upon the Delta method.<br>c. The sum-of-squared-residuals $V$ was computed as in equation (5). The Akaike information criterion ($AIC$) was computed as $2k + T \ln(V/T)$, where $k = 5$ parameters (including the parameter $\sigma^2$) and $T = 99$ observations. |  |  |

As indicated by the comparative values of the least squares criterion *V* and the corresponding Akaike information criterion (*AIC*) in Table 1, the model based upon all reported cases had a significantly tighter fit than the model based on cases of Omicron BA.1 alone. Overall, the Omicron BA.1-based model was only *exp*((*AIC*_*All Cases*_ − *AIC*_*BA*.1_)/2) = 0.0017 times as likely as the all cases-based model to minimize the information loss.

### 4.2. Plotting the Criterion V as a Function of the Parameters β and α

Figure 4A plots the least squares criterion *V* (blue curve, left axis) and the first partial derivative ∂*V*/∂*β* (red curve, right axis) as functions of the parameter *β*. The remaining parameters have been held constant at their estimated values, as shown in Table 1. The criterion *V* reached a minimum at the optimum *β*\* = 0.233, where ∂*V*/∂*β* = 0. The function *V* was convex in the interval from *β* = 0.169, where ∂*V*/∂*β* reached a minimum, to *β* = 0.352, where ∂*V*/∂*β* reached a maximum.

**Figure 4.**
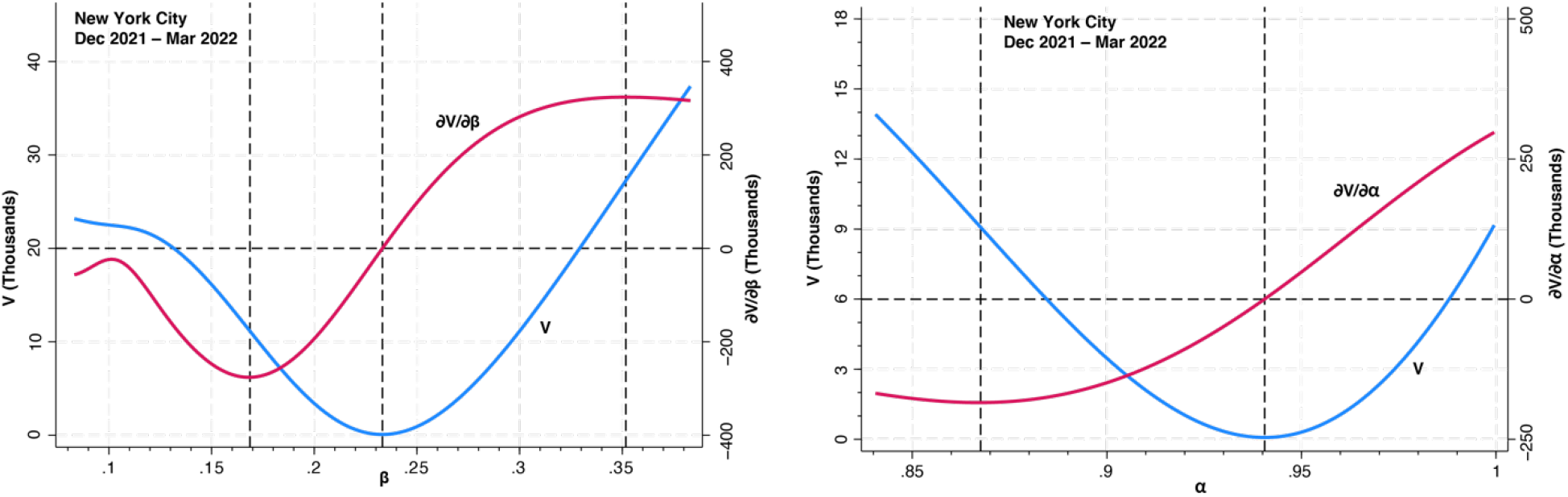
Panel A. Least Squares Criterion *V* (Left Axis) and First Partial Derivative *∂V*/*∂β* (Right Axis) as Functions of the Parameter *β*. Panel B. Least Squares Criterion *V* (Left Axis) and First Partial Derivative *∂D*/*∂α* (Right Axis) as Functions of the Parameter *α*.

Figure 4B displays the analogous plot of *V* and ∂*V*/∂*α* as functions of the parameter *α*, where the remaining parameters are similarly held constant at their optimum values. The criterion *V* reached a minimum at the optimum *α*\* = 0.941, at which point ∂*V*/∂*α* = 0. The function *V* was convex in the interval from *α* = 0.868, where ∂*V*/∂*α* reached a minimum, to *α* = 1, the boundary of admissible values of *α*, where ∂*V*/∂*α* remained positive.

Figure 5 plots the projection of *V* onto the (*β, α*) plane, once again based upon the data for all reported cases. As in Figure 4, the remaining parameters (*i*_0_, *N*) were held at the estimated values given in Table 1 above. The darkest area represents a ravine where *V* attained its lowest values. The yellow point in the center is the global minimum (*β*\*, *α*\*) = (0.233, 0.941).

**Figure 5.**
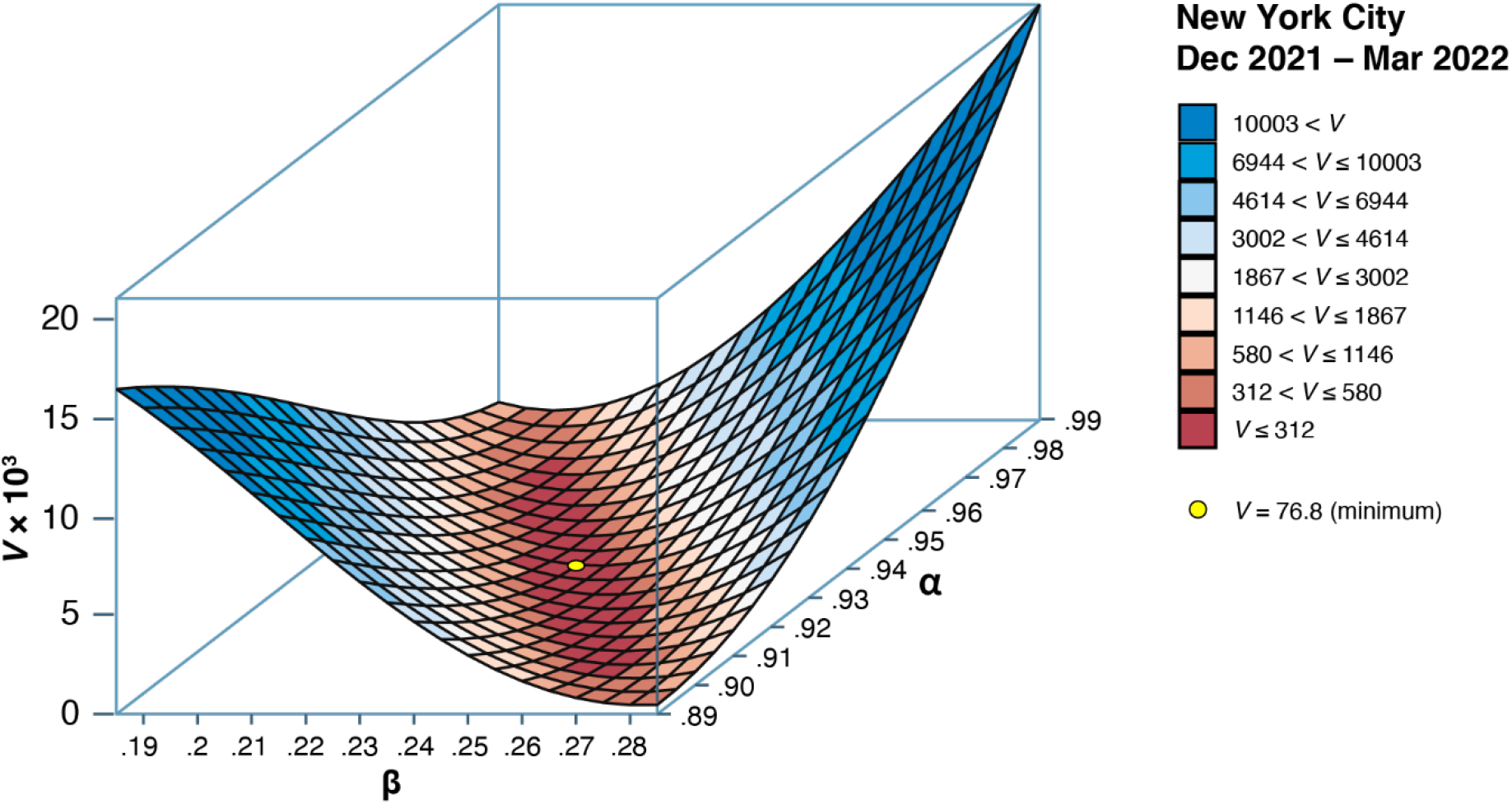
Least Squares Criterion *V* Projected onto the (*β, α*) Plane. The yellow point identifies the minimum where (*β*\*, *α*\*) = (0.233, 0.941). The parameters *i*_0_ and *N* were held constant at their optimum values given in Table 1. The plot was produced in part from the Stata program *surface* [72].

Figure 6 further characterizes the ravine identified in Figure 5. To that end, we relied on the results of Proposition 7 to identify the ray through the optimum point (*β*\*, *α*\*) along which the second-order directional derivative of *V* was minimized. Shown in the figure are the contours of *V* as a function of (*β, α*) running parallel to each of the two parameter axes, as well as the contour along the ray identified has having the flattest curvature. The ray corresponded to the equation *w*\*(*β* − *β*\*) + (*α* − *α*\*) = 0, where *w*\*= 1.073. The curvature along the ray, as measured by the second-order directional derivative, was less than one-tenth of the corresponding curvatures measured along the contours running parallel to the axes. Conditional on *w*\*, the 95% confidence interval surrounding the linear combination *w*\**β* + *α* had a very tight range of (1.183, 1.198).

**Figure 6.**
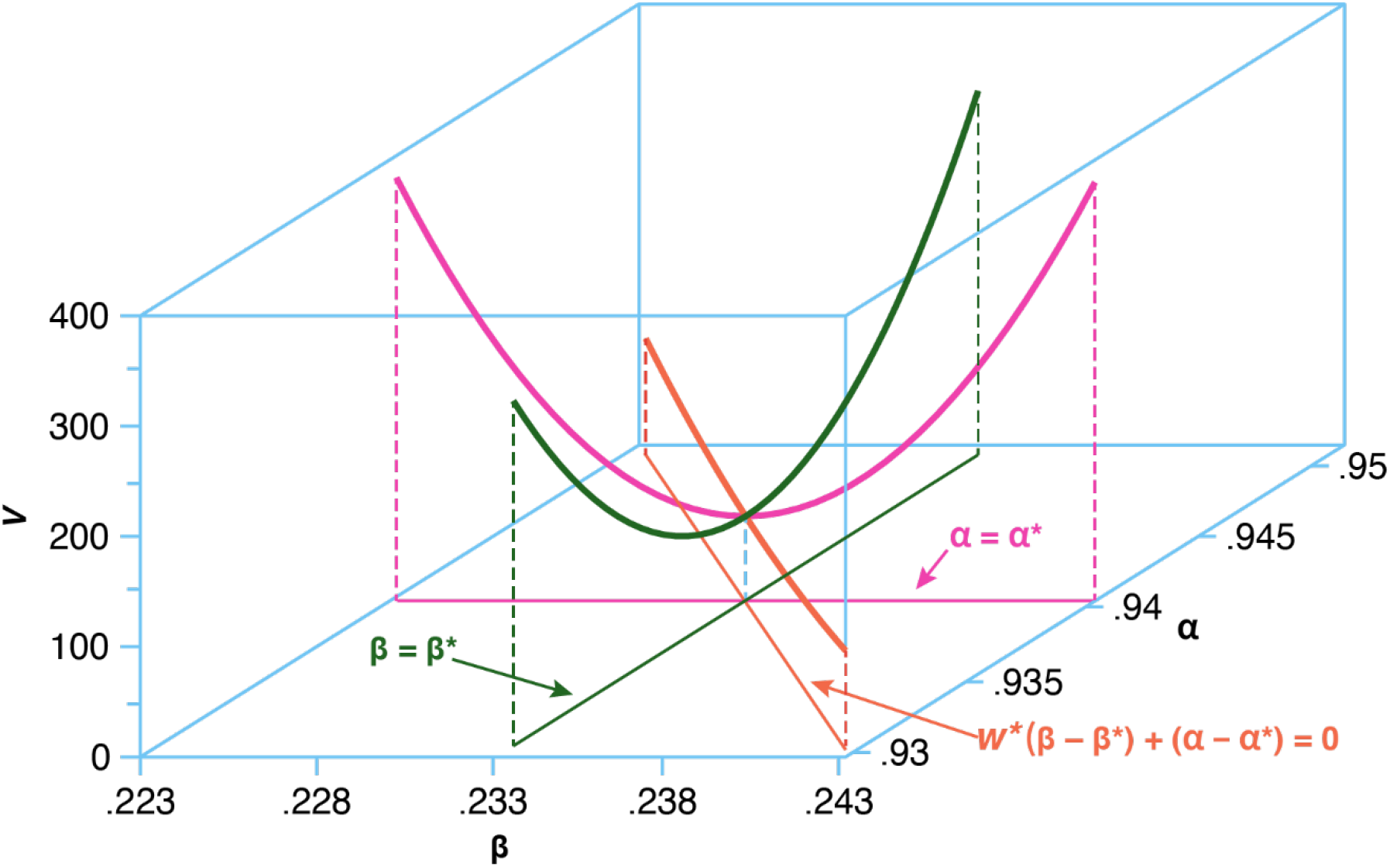
Contours of *V* Running through the Optimum Point Θ* Projected onto the (*β, α*) Plane. All plotted contours run through the optimum (*β*\*, *α*\*) = (0.233, 0.941), with the parameters *i*_0_ and *N* held constant at their optimum values. With *β* also held constant at its optimum value (green contour), the second-order derivative was ∂^2^*V*(***θ***\*)/∂*α*^2^ = 4.912×10^6^. With *αα* also held constant at its optimum value (magenta contour), the second-order derivative was ∂^2^*V*(***θ***\*)/∂*β*^2^ = 6.075×10^6^. Along the ray defined by *w*\* = 1.073 (orange contour), the second-order directional derivative was (*d*^2^*V*/*du*^2^)|_*u* =0_ = 0.418×10^6^.

In Appendix B, we further plot the path of successive iterations of the 4-parameter Newton-Raphson algorithm, showing how the estimate **Θ**^(*k*)^ entered the ravine at iteration *k* = 10 and then stopped at the optimum on iteration *k* = 40.

### 4.3. Consistency Checks with Data Simulated from Known Parameters

In Panel A of Figure 7, we plot the estimates *β*^(*j*)^ against the estimates *α*^(*j*)^ for *j* = 1, … 100 simulated epidemics with known parameters (*β, α, i*_*o*_, *N*) = (0.6, 0.7, 0.04, 10^5^) and thus with known basic reproduction number ℛ_0_ = *β*/(1 − *α*) = 2. The gold solid points represent the estimates based upon the 4-parameter Newton-Raphson algorithm, while the superimposed dark blue circles represent the estimates based on a 3-parameter Newton-Raphson algorithm conditional upon the population size *N* equal to its assumed value of 10^5^. In Panel B, we show the corresponding box-and-whisker plots of the estimated basic reproduction numbers 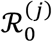 under the same two estimation conditions.

**Figure 7.**
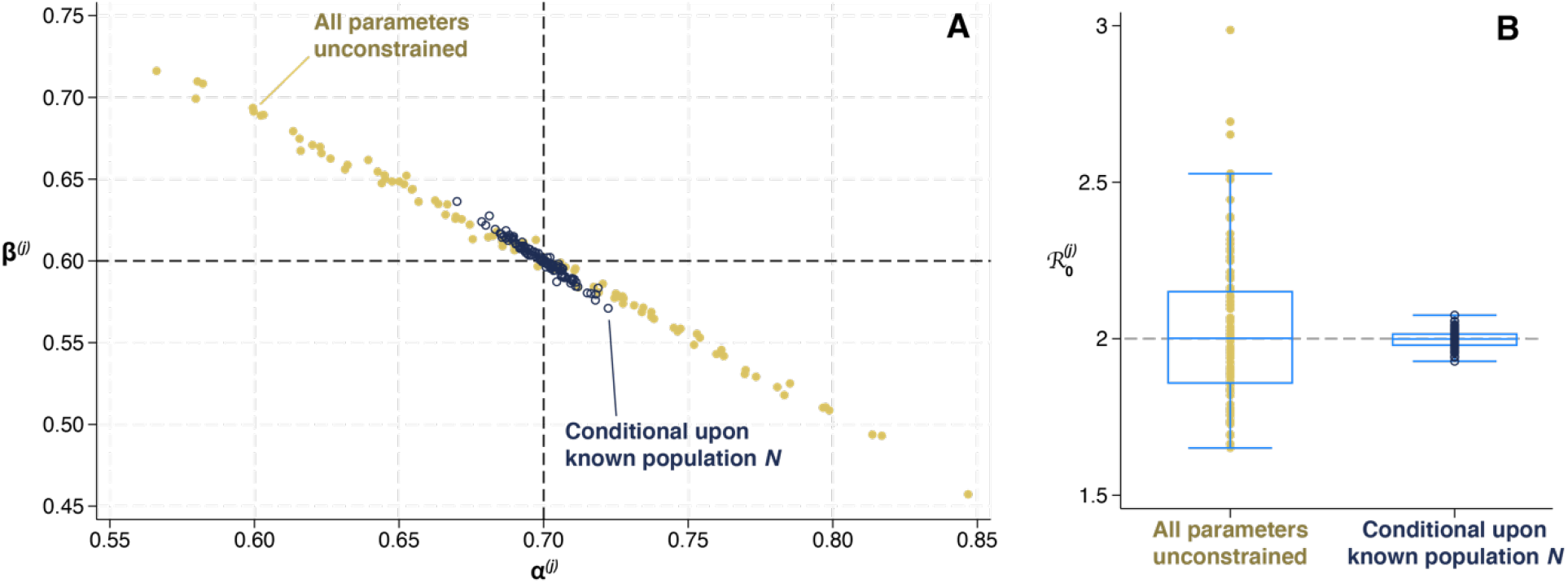
Estimated Parameters Based Upon a Simulated SIR Model with Gaussian Error. The SIR model of (2), (3) and (4) was simulated with assumed known parameters **Θ** = (*β, α, i*_*o*_, *N*) = (0.6, 0.7, 0.04, 10^5^) over *T* = 100 time periods. The observed case incidence was then computed as *y*_*t*_ = *X*_*t*_ + *ε*_*t*_, where the independent Gaussian errors *ε*_*t*_ were randomly drawn with zero mean and standard deviation *σ* = 100. Panel A. Plots of parameter estimates *β*^(*j*)^ versus *α*^(*j*)^ with all parameters unconstrained (gold solid points) and conditional upon a known population *NN* (dark blue circles). Panel B. Box-and-whisker plots of the corresponding estimates 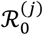 of the basic reproduction number. The boxes show the 25^th^, 50^th^, and 75^th^ percentiles while the capped lines show the bounds of interquartile range.

The plots in Figure 7 show that under both conditions, the estimates (*β*^(*j*)^, *α*^(*j*)^) were centered around their assumed values. Their respective sample means were (0.604, 0.696) with all four parameters unconstrained and (0.601, 0.699) conditional upon the known value of the population parameter *N*. The corresponding sample medians of (*β*^(*j*)^, *α*^(*j*)^) were (0.601, 0.699) and (0.600, 0.700) under the two respective estimation conditions. In addition, the sample means of the basic reproduction numbers 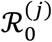 were 2.029 and 1.998 under the two respective estimation conditions, while their respective sample medians were 2.001 and 1.999.

Figure 7 confirms the strong negative correlation of the two estimated parameters, which were tightly situated along a near-linear paths. When we employed the 4-dimensional Newton-Raphson algorithm, the estimates were widely dispersed. Our conditioning upon the known value of the population parameter *N* substantially narrowed the dispersion, but it did not affect the observed strong negative correlation between the estimates of *β* and *α*. Thus, for the gold points in Figure 7A, representing the simulation results with unconstrained parameters, the ordinary least squares regression of *β*^(*j*)^ against *α*^(*j*)^ yielded a slope of −0.911 with 95% confidence interval [−0.924, −0.899]. For the dark blue points, representing the simulation results conditional on a known parameter *N*, the corresponding slope was −1.184 with 95% confidence interval [−1.225, −1.142].

### 4.4. Benchmarking

Estimation of the parameters **Θ** from the New York City Omicron data utilizing the 4-parameter Newton-Raphson method of Propositions 2 and 3 required 0.434 seconds of computational time, whereas estimation of **Θ** utilizing the EM-type method of Propositions 4 and 5 required 0.925 seconds. By contrast, estimation of the parameters utilizing an exhaustive grid search routine required 463.682 seconds, about three orders of magnitude longer than the 4-parameter Newton-Raphson routine.

## 5. Discussion

We have described a new strategy for estimating the underlying parameters of the Susceptible-Infected-Removed (SIR) model from observations on case incidence alone. Our approach finds the parameter vector that minimizes the sum of squared deviations between observed and predicted case incidence data. It does not require that we derive a closed-form mathematical expression for the predicted case incidence as a function of the parameters. Instead, we show how the gradient vector and the Hessian matrix of the least squares objective function can be exactly calculated. These quantities alone are sufficient to implement the Newton-Raphson algorithm to locate the parameter vector that minimizes the objective function.

### 5.1. Not All Five SIR Parameters Can be Identified from Case Incidence Data Alone

Our results shed light on the identifiability of the SIR model parameters from observations on case incidence alone. Our full SIR model had five parameters: *β*, the infection transmission parameter; *α*, the infection persistence parameter, gauging the proportion of infected individuals who remain in the infected state at each discrete time *t*; *i*_0_, the initial proportion of the population infected; *r*_0_, the initial proportion of the population in the removed state; and *N*, the total population.

In Proposition 1 of our theoretical analysis, we found that all five parameters were not jointly identifiable from the incidence data alone. (A related result appears to have been derived from a continuous-time SIR model with seasonal forcing [53].) Accordingly, we estimated the first four parameters under the identifying restriction that *r*_0_ = 0. In effect, we assumed that because of immune escape [73, 74], no one at the onset of the Omicron wave was already resistant to infection by the newly emergent SARS-CoV-2 variant.

### 5.2. Only a Linear Combination of β and α is Practically Identifiable

Our findings based upon the New York City Omicron data confirmed that the remaining four parameters **Θ** = (*β, α, i*_0_, *N*) could be indeed identified at a global interior minimum of the least squares objective function *V*. But we also uncovered a deeper problem of parameter identification. While the bivariate plots of Figure 4 demonstrated that the objective function was locally strictly convex at the optimum point, the three-dimensional plot of Figure 5 revealed that this point was situated along the floor of a ravine where the parameters *β* and *α* were highly correlated. In Figure 6, we were able to identify the floor of the ravine as the line in the (*β, α*) plane where the second-order derivative of *V* was minimized. While the individual parameters *β* and *α* had wide estimated confidence intervals, the linear combination of *β* and *α* along the ravine could be tightly estimated. Appendix Figure B1 further showed how successive estimates **Θ**^(*k*)^ of the four-dimensional Newton-Raphson algorithm followed along the ravine floor until the point of convergence.

What’s more, our consistency checks on a simulated database confirmed that the extremely high correlation between the parameters *β* and *α* was not an idiosyncrasy of the New York City Omicron data. The results shown in Figure 7, based on simulated data, demonstrated that while the dispersion of the estimates *β*^(*j*)^, *α*^(*j*)^ were centered around their true values, they similarly tended to demonstrate a near-linear relation between the two parameters. Conditioning on the population size parameter *N* reduced the dispersion of the estimates in the (*β, α*) plane as well as the estimated basic reproduction number ℛ_0_ – a finding consistent with the narrower confidence intervals shown in Table 1 for the New York City Omicron data – but it did not attenuate the strong negative correlation between the parameters.

Even with the identifying restriction that the initial proportion of recovered individuals *r*_0_ is zero, the remaining four-parameter version of SIR is what some physicists have described as a “sloppy” model [52, 54]. While *β* and *α* are, strictly speaking, separately identifiable from case incidence data, we conclude that only a linear combination of the two parameters is *practically* identifiable [66]. (A related result appears to have been derived in [75].)

### 5.3. We Cannot Rely on Census Data Alone to Identify the Parameter N

Imposing the strong identifying assumption that the initial proportion *r*_0_ of recovered individuals was zero, we estimated the population size parameter *N* to be 1.013 million, which amounted to only 11.9 percent of New York City’s total census population of 8.5 million. This discordant result suggests that it might have been more appropriate to set *N* = 8.5 million as the identifying assumption. In that case, however, relying upon Proposition 1, we would then end up with a rather large estimate of *r*_0_ = 0.881, which would in turn imply 1 − *r*_0_ = 0.119 and thus *N*(1 − *r*_0_) = 1.013 million. Despite the evidence of immune escape on the part of the Omicron variant [74], we would conclude that a very substantial fraction of the population still retained long-term cellular immunity, possibly through the administration of multiple vaccine doses [76].

Unfortunately, such an alternative strategy of relying on census-based population data to identify the parameter *N* runs into two important complications: underreporting and incomplete mixing. In many contexts, particularly in recent applications of the SIR and related compartmental models to COVID-19 incidence, it has been widely recognized that a significant number of incident cases may have gone unreported [77]. There is concrete evidence of significant underreporting of Omicron infections, due principally to the widespread availability of home rapid antigen testing [77], particularly in New York City [78, 79]. In the absence of reliable information on the temporal pattern of such underreporting, the most parsimonious approach to this phenomenon has been to assume a *case identification ratio* equal to a constant *p* < 1 [75, 80, 81]. In that case, our model would need to be modified to accommodate the reality that our reported incidence data *y*_*t*_ are in fact estimates of *pX*_*t*_ rather than *X*_*t*_. We’ve learned from Proposition 4, however, that each output variable *X*_*t*_ can be written in the form *φ*_*t*_(***θ***)*N*, where *φ*_*t*_(***θ***) is a function of the remaining identifiable parameters ***θ*** = (*β, α, i*_0_). Accordingly, our reported incidence data *y*_*t*_ are really estimates of *pX*_*t*_ = *φ*_*t*_(***θ***)(*pN*). To identify our model, we thus need a prior estimate of *pN*. Unless we’re prepared to impose additional restrictions on *p*, census data on *N* alone will not be enough.

We would ordinarily interpret the parameter *N* to gauge the size of the population at risk for contagion. This population would consist of all individuals who homogeneously mix with each other in accordance with the law of mass action embodied in equations (1) and (3). Many investigators, however, have properly recognized that the underlying assumption of homogeneous mixing may not apply to the entire population [19, 82-87]. This caution applies just as well to the Omicron wave, when a substantial proportion of the population avoided retail establishments, drinking and eating places, transportation venues, worksites and other high-risk locations [88].

Take the case of the COVID-19 outbreak at the campus of the University of Wisconsin-Madison in September 2020, where total student enrollment was 44, 640, but where the large fraction of cases was concentrated in two on-campus student residence halls with a combined population of about 2, 932 [89]. If we relied on the campus-wide census to identify the parameter *N* on the campus-wide population, we would have ended up concluding erroneously that about 94 percent of the student population was already immune to SARS-CoV-2.

### 5.4. Should We Exclude the Non-Omicron Case Counts from our Database?

In Figure 3, we saw that exclusion of non-Omicron-variant cases from our database resulted in a fitted model that overshot the initial upswing of cases during December 2021. That is, the predicted output variable *X*_*t*_ exceeded the observed counts 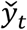 during *t* = 1, …, 10. Our AIC computations in Table 1 confirmed that dropping the residual Delta-variant infections from the previous wave significantly reduced the overall goodness of fit of our SIR model to the new Omicron wave data.

Despite the evidence favoring immune escape by Omicron [74], these residual Delta-variant infections may have conferred partial cross-immunity against the newly emerging Omicron variant and thus reduced the initial proportion of individuals susceptible to Omicron BA.1 [90]. If so, excluding these Delta cases would upwardly bias the model estimates of the numbers of susceptible individuals *S*_*t*_ and, consequently, the predicted output variable *X*_*t*_.

Still, the apparent preference for inclusion of the non-Omicron cases is hardly clear cut. The unconditional confidence intervals are simply too wide to distinguish sharply between the corresponding parameter estimates in the two columns of Table 1.

### 5.5. Our Estimates of the Parameter α Appear to be Inconsistent with Clinical Observation

Our estimates of the parameter *α* appear to be out of line with what is known from direct clinical measurement about the duration of infectivity from the Omicron variant. In one cohort of 55 symptomatic Omicron-infected patients, only 13.5 percent continued to shed virus ten days after infection [91]. Yet our point estimates of the proportion remaining infectious after ten days (computed as *α*^10^) would be 54.4 percent from the combined-variant data and 36.0 percent from the Omicron-only data. A meta-analysis of 29 clinical studies suggested that while the mean duration of polymerase chain reaction (PCR) positivity was 10.82 days, the mean duration of viral shedding was only 5.16 days [92]. Yet our point estimates of the mean duration of infectivity (computed as 1/(1 − *α*)) were 16.8 days from the combined-variant data and 10.3 days from the Omicron-only data.

We need to interpret this apparent discrepancy with caution. The unconditional confidence intervals around our estimates of *α* were quite wide. A reported short mean duration of infectivity does not rule out a long-tailed distribution. In one study, 25 percent of patients were still shedding virus after 8 days [93]. What’s more, an individual’s infectivity is not an all-or-nothing characteristic. It is likely to depend on the extent to which an infected individual isolates himself from others. That behavioral component of the parameter *α* may have changed during the Omicron wave.

### 5.6. Is SIR the Correct Structural Model for Omicron?

Despite its close fit to the data in Figure 3, the Susceptible-Infected-Recovered framework may thus fail as a *structural* model of the Omicron wave, even if it apparently succeeds as a *reduced form* model [94]. When we imposed an identifying restriction on the parameter *r*_0_ representing the initial proportion of recovered individuals, we could indeed back out the remaining parameters (*β, α, i*_0_, *N*) from the observed case incidence data and thus solve the inversion problem. But the resulting parameter estimates do not necessarily warrant the *structural* interpretation that we assumed in our exposition of the SIR model in equations (1) through (5) above.

That is not to say, however, that an adequate reduced form model is incapable of making accurate projections [95]. But when it comes to sharply identifying individual parameters and thus distinguishing between alternative epidemic models, some prior information above and beyond case incidence data may be required.

### 5.7. Other Limitations and Extensions

Parameter search algorithms such as Newton-Raphson [46] have superior computational performance when they rely on exact expressions for the gradient vector and Hessian matrix of second derivatives, rather than on numerical approximation or brute-force search [45]. Still, as Figure 4 shows, there may be regions of the parameter space where the least squares criterion function is non-convex and the algorithm does not converge. While Appendix B displays convergence from a suitably chosen starting point, the tasks of finding the right initial values and keeping the parameter search within bounds remain unavoidable challenges.

In a benchmarking study of the New York City Omicron data, our four-parameter Newton-Raphson routine converged approximately 500 times faster than exhaustive grid search. (See section 4.4.) While Newton-Raphson is known to have quadratic convergence so long as the initial conditionas are correctly specified, additional research will be required to compare the efficiency of our approach with other estimation algorithms, especially as the sample size varies.

To approximate a continuous-time dynamic model as closely as possible, we relied on daily counts of reported COVID-19 cases. As Figure 1 shows, however, such a fine level of detail resulted in striking day-of-the-week effects, which required us to pretreat the raw case counts. Our centered, 7-day moving average appeared to be the most flexible nonparametric approach to this task. Alternative approaches that incorporate parametric models of day-of-the-week effects directly in our model of the output variable *X*_*t*_ have yet to be tried.

Applying our approach to the New York City Omicron wave, we have estimated SIR model parameters from counts of reported incident cases. Our approach can be generalized to estimate SIR model parameters from counts of removed cases, either through recovery or death [96].

We have focused sharply on the original SIR model, rather than its numerous variations. Still, our basic approach can be extended to these more complex models. Our findings offer a caution, however, that such models as SEIR, which require an additional parameter governing the transition from an intermediate *exposed* state to the *infected* state, may very well turn out to be sloppy [75].

One exemption may be the well-studied SIRS model [58, 97], where individuals in the *R* (recovered) state can transition back to the *S* (susceptible) state due to waning immunity. The SIRS model still has three states, but the parameter vector has an additional component governing the rate of transition from *R* back to *S*. Since the SIRS model is known to admit oscillations with a stable endemic equilibrium [58, 98], we would be in a position to run our parameter recovery algorithm on multiple waves of data *y*_*t*_. That is a task for future research.

## Data Availability

The data and programs used in this work are publicly available on the Open Science Framework at https://osf.io/3b4hv/.

## 6. Appendix A. Proofs of Propositions

### Proposition 1.

*Proof*. We characterize the mapping *ϕ* from the five-parameter vector **Θ** into a four-parameter vector *ϕ*(**Θ**) = (*κ, λ, µ, ν*) as follows:

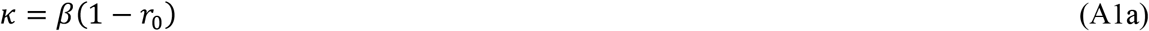

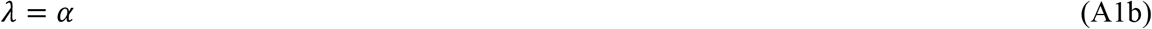

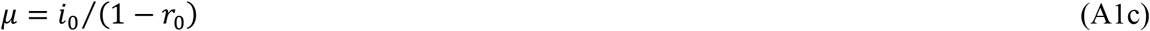

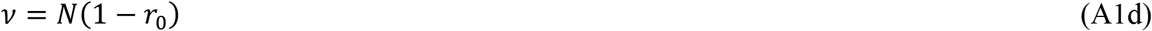

Now consider two distinct parameter vectors **Θ** = (*β*, *α, i* _0_, *r* _0_, *N*) and 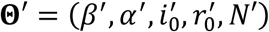, where **Θ** ≠ **Θ**′. The condition *ϕ* (**Θ**) = *ϕ* (**Θ** ′) requires that:

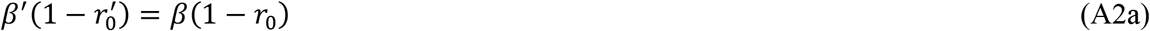

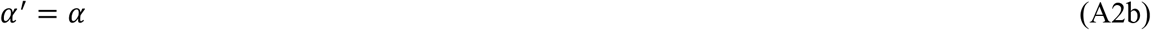

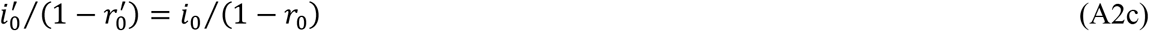

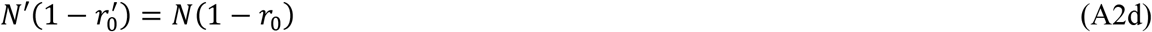

We need to show that ***X***(**Θ**) = ***X***(**Θ**′). From (3) in the main text, each component *X*_*t*_ of the vector ***X***(**Θ**) in turn depends on the ratio (*β*/*N*) as well as the corresponding time-specific state variables *S*_*t*_ and *I*_*t*_. Dividing (A2a) by (A2d), we see that (*β*/*N*) = (*β*′/*N*′). Hence, it remains only to show that *S*_*t*_(**Θ**) = *S*_*t*_(**Θ**′) and *I*_*t*_(**Θ**) = *I*_*t*_(**Θ**′) for all times *t* = 0, …, *T*.

We proceed by mathematical induction. First consider time *t* = 0. For the respective parameter vectors **Θ** and **Θ**′, we have *S*_0_(**Θ**) = (1 − *i*_0_ − *r*_0_)*N* and 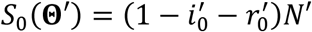. From (A2c), we can write 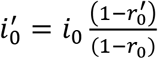, and from (A2d), we can write 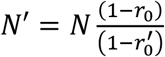. So, 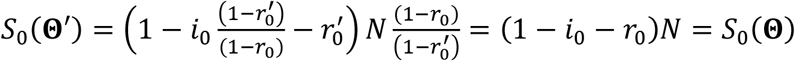. Similarly, we have *I*_0_(**Θ**) = *i*_0_*N* and 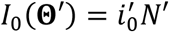. Again, it follows from (A2c) and (A2d) that 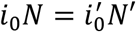 and thus *I*_0_(**Θ**) = *I*_0_(**Θ**′). So, the proposition holds for *t* = 0.

Now assume that *S*_*t*_(**Θ**) = *S*_*t*_(**Θ**′) and *S*_*t*_(**Θ**) = *I*_*t*_(**Θ**′) for any *t* ≥ 0. We show that *S*_*t*+1_(**Θ**) = *S*_*t*+1_(**Θ**′) and *I*_*t*+1_(**Θ**) = *I*_*t*+1_(**Θ**′). From (1a) in the main text, we have *S*_*t*+1_(**Θ**) = *S*_*t*_(**Θ**) − *βS*_*t*_(**Θ**)*I*_*t*_(**Θ**)/*N*. We’ve already learned from (A2a) and (A2d) that (*β*/*N*) = (*β*′/*N*′), so we can instead write *S*_*t*+1_(**Θ**) = *S*_*t*_(**Θ**) − *β*′*S*_*t*_(**Θ**)*I*_*t*_(**Θ**)/*N*′. Since we’ve assumed that *S*_*t*_(**Θ**) = *S*_*t*_(**Θ**′) and *S*_*t*_(**Θ**) = *I*_*t*_(**Θ**′), we can therefore write *S*_*t*+1_(**Θ**) = *S*_*t*_(**Θ**′) − *β*′*S*_*t*_(**Θ**′)*I*_*t*_(**Θ**′)/*N*′ = *S*_*t*_(**Θ**′). Similarly, from (1b), we have *I*_*t*+1_(**Θ**) = *βS*_*t*_(**Θ**)*I*_*t*_(**Θ**)/*N* + *αI*_*t*_(**Θ**). Substituting (*β*/*N*) = (*β*′/*N*′) and *α*′ = *α* from equation (A2b) gives *I*_*t*+1_(**Θ**) = *I*_*t*+1_(**Θ**′). ▪

### Corollary 1

The four components of the parameter vector **Φ** = (*κ, λ, µ, ν*) in the following SIR model can be identified from the data ***y*** on case incidence:

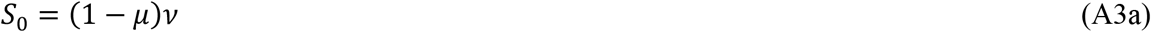

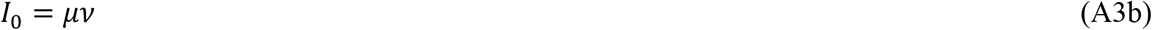

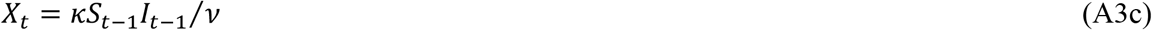

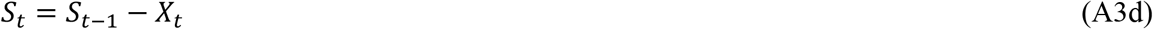

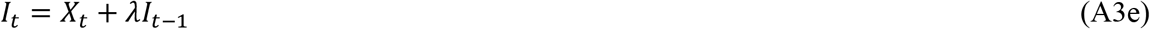

*Proof*. We repeat the basic equations of our dynamic model, as articulated in equations (1), (2), and (3) of the main text:

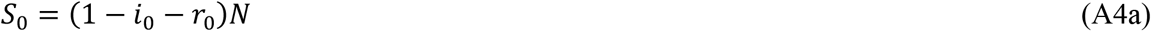

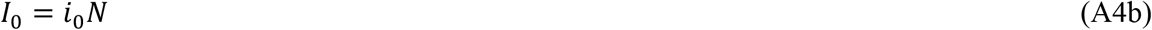

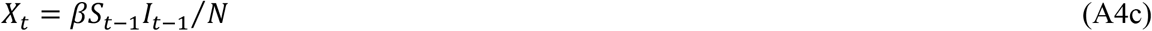

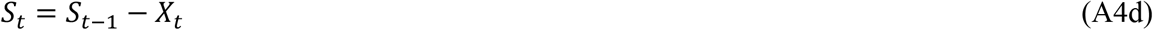

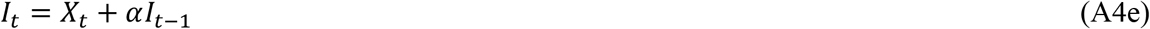

Now substitute definitions of (*κ, λ, µ, ν*) given in equations (A1) in the proof of Proposition 1.

To define the new four-dimensional parameter vector **Φ**, we have essentially dropped the parameter *r*_0_ from the original five-dimensional vector **Θ** and then normalized the remaining four parameters (*β, α, i*_0_, *N*) by the factor (1 − *r*_0_). The resulting mapping ***X***(**Φ**), as defined in (A4) is one-to-one. ▪

### Corollary 2

The sharp prior restriction that *r*_0_ = 0 is sufficient to identify the remaining four parameters (*β, α, i*_0_, *N*) of the five-parameter vector **Θ**.

*Proof*. Consider a specific parameter vector **Θ** = (*β, α, i*_0_, 0, *N*) with *r*_0_ = 0. From equations (A2) in Proposition 1, any other parameter vector 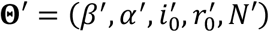 satisfying *ϕ*(**Θ**′) = *ϕ*(**Θ**) and therefore ***X***(**Θ**′) = ***X***(**Θ**) must satisfy these conditions:

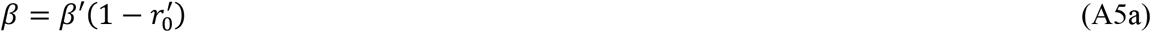

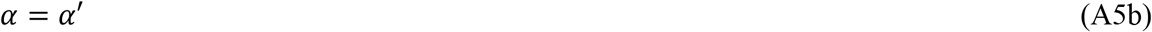

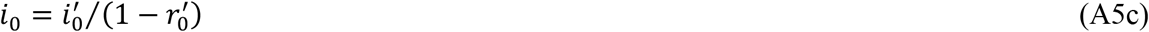

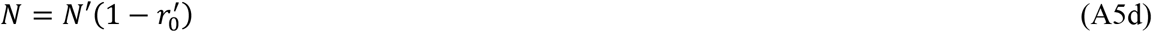

However, the only vector 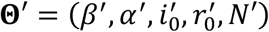 satisfying these conditions *and* adhering to the restriction that *r*_0_^′^ = 0 is the vector **Θ** itself. Hence, **Θ** is uniquely identified. We further note that under the restriction that *r*_0_ = 0, the vector of the remaining four parameters (*β, α, i*_0_, *N*) is exactly equal to the four-parameter vector **Φ** = (*κ, λ, µ, ν*) defined in Corollary 1. ▪

### Corollary 3

A sharp prior restriction on any one of the four parameters (*β, i*_0_, *r*_0_, *N*) is sufficient to identify the full five-parameter vector **Θ** = (*β, α, i*_0_, *r*_0_, *N*). A sharp prior restriction on the parameter *α* alone, however, will not identify the remaining parameters.

*Proof*. We’re essentially implementing the proof strategy for Corollary 2. For example, let’s impose the sharp prior restriction that the population size *N* is known and equal to 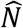, and consider a specific parameter vector 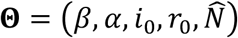 with 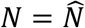. Any other parameter vector 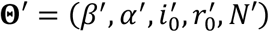 satisfying *ϕ*(**Θ**′) = *ϕ*(**Θ**) and therefore ***X***(**Θ**′) = ***X***(**Θ**) must satisfy the conditions in equations (A2) in Proposition 1 as well as the additional restriction that 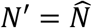. Since equation (A2d) alone requires that 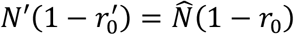, the additional restriction would imply that 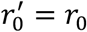. From the remaining equations in (A2), we therefore conclude that *β*′ = *β, α*′ = *α*, and 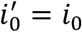 as well. Hence, **Θ** is uniquely identified. We note that imposing a prior restriction that the parameter *α* alone does no more than repeat the requirement of equation (A2b) and hence does not identify the remaining parameters. However, a sharp prior restriction on the basic reproductive number ℛ_0_ would also be sufficient to identify all five parameters of the vector **Θ**. ▪

### Proposition 2.

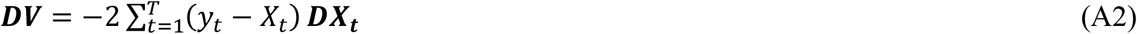

where each column vector 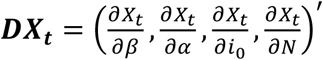 represents the corresponding gradient of partial derivatives of the output variable *X*_*t*_ at time *t*. The basic equations of our dynamic system (1), in combination with the initial conditions (2), can be used to generate complete, computable difference equations for ***DX***_***t***_ for all *t*, and thus for ***DV***.

*Proof*. Equation (A2), which repeats equation (6), is derived by taking the derivative of *V* as defined in equation (5). Taking the derivative of ***DX***_***t***_, we get:

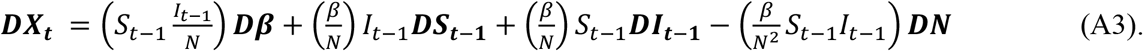

where ***Dβ*** = (1, 0, 0, 0)′ and ***DN*** = (0, 0, 0, 1)′. Taking the derivative of *S*_*t*_ as defined in equation (1a), we get:

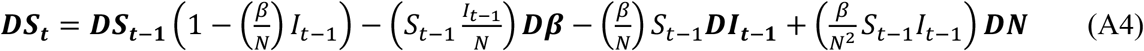

From equations (1b) and (3), we can write 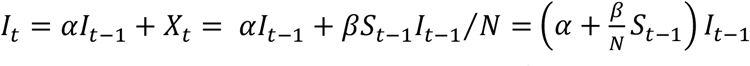. Taking the derivative of this expression gives:

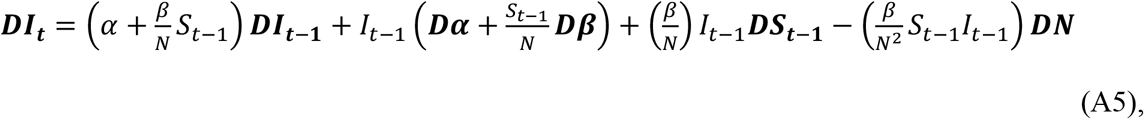

where ***Dα*** similarly represents the unit vector (0, 1, 0, 0) ′.

To complete the calculation, we need to compute the initial values of the gradients ***DS***_**0**_ and ***DI***_**0**_. Since *I*_0_ = *i*_0_*N* and *S*_0_ = (1 − *i*_0_)*N*, we can write the gradients ***DI***_**0**_ = *N****Di*** _**0**_ + *i*_0_***D*** and ***DS***_**0**_ = −*N****Di***_**0**_ + (1 − *i*_0_)***D***, where ***Di***_**0**_ = (0, 0, 1, 0)′. Given these initial gradient values, we can use (A4) and (A5) to iteratively compute the vectors ***DS***_***t***_ and ***DI***_***t***_ for all *t* = 1, …, *T*. Once vectors ***DS***_***t***_ and ***DI***_***t***_ have been computed, we can then apply (A3) to iteratively compute the corresponding gradients ***DX***_***t***_. Those quantities in turn yield the gradient ***DV*** of our objective function through equation (A2). ▪

### Proposition 3.

Let ***DX*** denote the *T* × 4 matrix whose *t*-th row is the vector 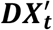, as defined in Proposition 2. The Hessian matrix of the least squares criterion *V* with respect to the parameter vector **Θ** = (*β, α, i* _0_, *N*) is:

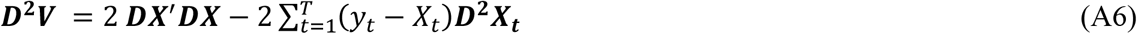

*Proof*. To compute the Hessian matrix ***D***^**2**^***V***, we need to determine the corresponding Hessian matrices ***D***^**2**^***X***_***t***_ for all *t*. To that end, we adopt the following notation. For two column vectors ***A*** and ***B*** of dimension *L* × 1, we define the *L* × *L* symmetric matrix ***A***⨀***B*** = ***AB***′ **+ *BA***′. Now taking the derivative of ***DX***_***t***_ in (A3), we get:

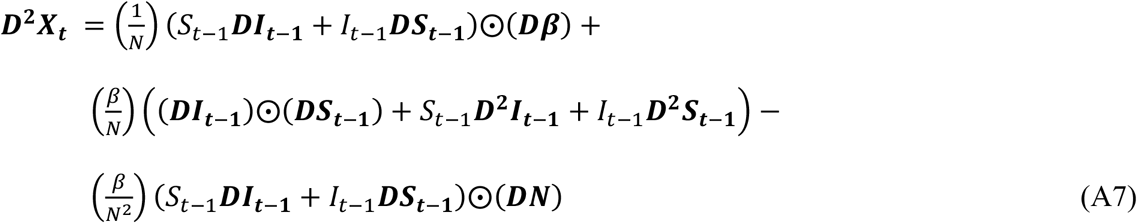

Thus, the Hessian matrices ***D***^**2**^***X***_***t***_ depend in turn on the lagged gradient terms ***DS***_***t***−**1**_ and ***DI***_***t***−**1**_ as well as the lagged Hessian matrices ***D***^**2**^***S***_***t***−**1**_ and ***D***^**2**^***I***_***t***−**1**_. The corresponding expressions for the latter Hessian matrices are:

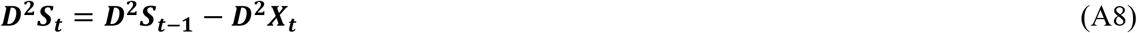

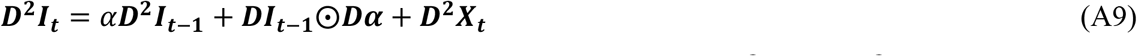

To complete our calculations, we note that the initial values ***D***^**2**^***S***_**0**_ and ***D***^**2**^***I***_**0**_ are simply null atrices.

Taken together, equations (A6), (A7), (A8) and (A9), along with the initial null values of ***D***^**2**^***S***_**0**_ and ***D***^**2**^***I***_**0**_, permit us to iteratively compute the Hessian ***D***^**2**^***V***, just as we outlined for the gradient ***D*** in the proof of Proposition 2 above. ▪

### Application to Aggregate Data on Case Incidence

In some applications, we may have only aggregate data reported over a coarse time scale. For concreteness, let’s assume that the underlying SIR epidemic model is valid when the time axis *t* =0, 1, …, *T* is marked off in days, but we only have data on the cumulative number of cases during each 7-day week, indexed by *m* = 1, …, *M*. Days *t* are mapped into weeks *m* by the relation 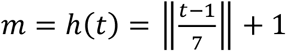, where the floor operator ‖*x*‖ maps into the largest integer *j* such that *j* ≤ *x*. We further assume that *T* = 7*M*, so that week *M*, which corresponds to the last week, ends on day *T*.

Now define the *M* × *T* aggregation matrix ***W*** with element *w*_*mt*_ = 1 if and only if *m* = *h*(*t*). Otherwise, *w*_*mt*_ = 0. While our disaggregated model generates daily values *X*_*t*_ of the latent variable from the underlying parameters ***θ***, our data *y*_*m*_ represent observations on the aggregated values 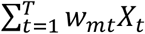. In vector notation, our least squares criterion would be *V* = (***y*** − ***WX***)′(***y*** − ***WX***). The corresponding gradient would be ***DV*** = −2 (***y*** − ***WX***)′***WDX***. The corresponding Hessian matrix would be ***D***^**2**^***V*** = −2 (***y*** − ***WX***)′***WD***^**2**^***X*** + 2(***WDX***) ∙ (***WDX***). With those modifications, the foregoing results in Propositions 2 and 3 would apply. For further details, see [96].

#### Proposition 4.

*Proof*. To prove this result, we only need to show that the state variables *S*_*t*_ and *I*_*t*_ are themselves proportional to *N*. Thus, if for all times *t* = 1, …, *T*, we can show that *S*_*t*_ = *f*_*t*_(***θ***)*N* and *I*_*t*_ = *g*_*t*_(***θ***)*N* for some functions *f*_*t*_(***θ***) and *g*_*t*_(***θ***), then from equation (3), we would have *X*_*t*_ = *βS*_*t*−1_*I*_*t*−1_/*N* = *βf*_*t*−1_(***θ***)*g*_*t*−1_(***θ***)*N*. Thus, *φ*_*t*_(***θ***) = *βf*_*t*−1_(***θ***)*g*_*t*−1_(***θ***).

We can prove that *S*_*t*_ and *I*_*t*_ are proportional to *N* by mathematical induction. From equations (2a) and (2b), respectively, we known that *S*_0_ = (1 − *i*_0_)*N* and *I*_0_ = *i*_0_*N*. So, the proposition is true for *t* = 0. Now suppose that *S*_*t*_ = *f*_*t*_(***θ***)*N* and *I*_*t*_ = *g*_*t*_(***θ***)*N*, at time *t*. We claim that *S*_*t*+1_ = *f*_*t*+1_(***θ***)*N* and *I*_*t*+1_ = *g*_*t*+1_(***θ***)*N* necessarily hold for some functions *f*_*t*+1_(***θ***) and *g*_*t*+1_ (***θ***). From (1a), we can write 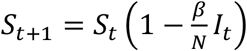. We have 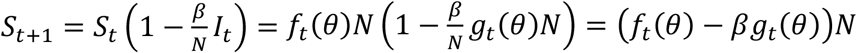, and so *f*_*t*+1_ (***θ***) = *f*_*t*_ (***θ***) − *βg*_*t*_ (***θ***). Similarly, 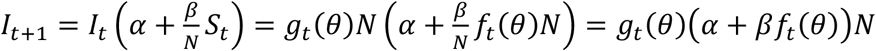, and so *g*_*t*+1_(*θ*) = *g*_*t*_(*θ*)(*α* + *βf*_*t*_(***θ***)). ▪

#### Proposition 5.

Let **Θ**^(*n*)^ = ***θ***^(*n*)^, *N*^(*n*)^ denote parameter estimates at iteration *n* of an iterative estimation algorithm. Let ***X***^(***n***)^ denote the corresponding output variable vector derived from the SIR model (2)-(4) based upon these parameter estimates. Define

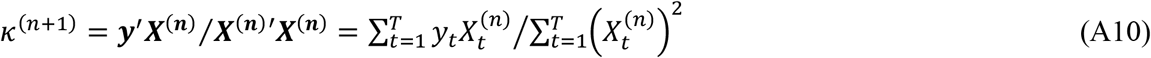

*Proof*. From Proposition 4, we know that each output variable *X*_*t*_ can be written in the form *φ*_*t*_(***θ***)*N*, where *φ*_*t*_(***θ***) is a function of the remaining parameters ***θ*** = (*β, α, i*_0_). Substituting into equation (5) in the main text gives:

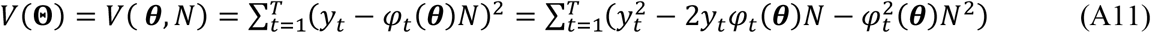

Equation (A11), which holds for all **Θ** = (***θ***, *N*), teaches us that the least squares criterion *V* is a quadratic function of the population-size parameter *N*.

Adhering to the assumptions of the proposition, let’s assume that we have parameter estimates **Θ**^(*n*)^ = ***θ***^(*n*)^, *N*^(*n*)^ at iteration *n* of an iterative procedure. Conditional on ***θ***^(*n*)^, the first-order condition for minimizing *V*(***θ***^(*n*)^, *N*) as a function of *N* is:

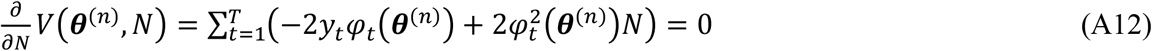

The unique root is:

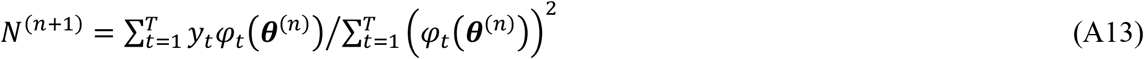

From Proposition 4, we have 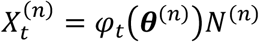. Substituting into (A13) and rearranging gives:

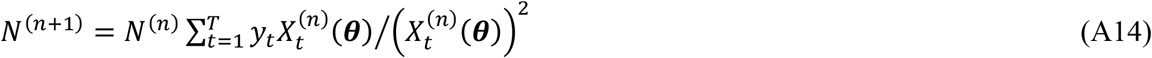

We have *N*^(*n*+1)^ = *κ*^(*n*+1)^*N*^(*n*)^, where *κ*^(*n*+1)^ is defined in (A10). ▪

#### Proposition 6.

Let 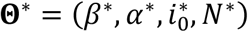 and denote the estimated parameter values that minimize the objective function *V*(**Θ**) under the identifying restriction *r*_0_ = 0, and let *V*\* = *V*(**Θ**\*) denote the corresponding minimized value of the least squares criterion *V*. Then the variance-covariance matrix of the estimated parameters **Θ**\* is ***C***\* = (*s*\*)^2^(***DX***\*′***DX***\*)^−1^, where (*s*\*)^2^ = *V*\*/*T*, and where ***DX***\*, as defined in Proposition 3, is also evaluated at the optimum **Θ**\*.

*Proof*. This result, which follows directly from the general statistical treatment of nonlinear least squares in [47], gives us the *unconditional* 4 × 4 variance-covariance matrix of the parameter vector 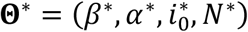 estimated from our 4-dimensional Newton-Raphson algorithm described in Propositions 2 and 3. If we instead employ the EM-type algorithm characterized in Propositions 4 and 5, then at the M-step of the final stage, we can obtain the 3 × 3 variance-covariance matrix of the parameter vector 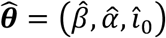 *conditional* upon 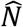, which has already been estimated at the E-step. In that case, we obtain 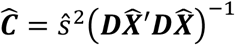, where 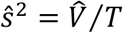, where 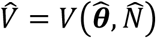, where and 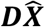 is the *T* × 3 matrix whose *t*-th row is the gradient of *X*_*t*_ with respect to ***θ*** evaluated at the optimum 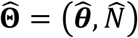. Since both the 4-dimensional Newton-Raphson algorithm and EM-type algorithm result in parameter estimates that minimize the objective function *V*(**Θ**), that is, since 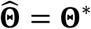, we can readily compute 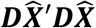 as the 3 × 3 submatrix of ***DX***\*′***DX***\*. ▪

#### Proposition 7.

Let **Θ**\* be a local interior minimum of *V*(**Θ**). Define a single-valued mapping from the real line into the four-dimensional subspace Ω of admissible values of **Θ** as follows: 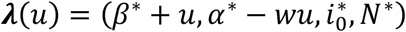, where *u* is its real-valued argument and *w* > 0 is a positive real parameter. As the parameter *w* varies, this mapping characterizes a family of rays in four-dimensional space passing through the point **Θ**\* at *u* = 0. Then there exists a ray, corresponding to a specific value of *w*, along which the second-order directional derivative 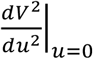 is nearly zero, that is, along which *V*(**Θ**) is *nearly* equal to its local minimum value *V*(**Θ**\*).

*Proof*. Along any such ray, defined by a specific value of the parameter *w*, the linear combination *wβ* + *α* is constant and equal to *wβ*\* + *α*\*, independent of the value of *u*. Moreover, for all *w*, we have ***λ***(0) = **Θ**\* and the composition *V* ∘ ***λ***(0) = *V****λ***(0) = **Θ**\*. For any specific value of *w*, the derivative of ***λ***(*u*) with respect to *u* is ***Dλ*** = (1, −*w*, 0, 0), which is the directional vector of the ray. Accordingly, the directional derivative of *V* along the ray ***λ***(*u*) at the minimum **Θ**\* is the inner product ***Dλ DV***(**Θ**\*), which simplifies to the expression 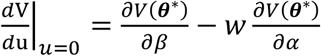. Moreover, the second-order directional derivative is the quadratic form ***Dλ D***^**2**^***V***(**Θ**\*) ***Dλ***′, which similarly simplifies to:

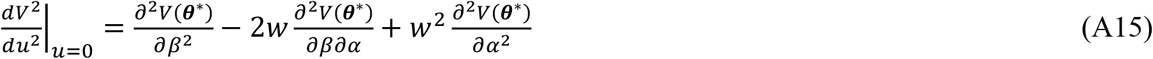

Our objective function *V* is the flattest along the ray for which this second-order directional derivative is minimized. From (A15), we see that the second-order directional derivative is itself a quadratic function of the parameter *w*. This quadratic function in turn reaches a minimum at 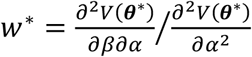. Plugging this value into (A15) gives the second-order directional derivative 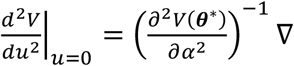, where ∇ is the determinant of the 2 × 2 submatrix of ***D***^**2**^***V***(**Θ**\*) corresponding to the parameters (*β, α*). As we have seen in the empirical implementation, this determinant is positive but quite small. ▪

## 7. Appendix B. Path of the 4-Parameter Newton-Raphson Algorithm: New York City Data

Figure B1 displays the path of each successive iteration ***θ***^(***k***)^ generated by the 4-dimensional Newton-Raphson algorithm, along with the corresponding values ***V***^(***k***)^ of the criterion function. As in Figures 5 and 6, we plot only the two parameter coordinates *β*^(*k*)^, *α*^(*k*)^, but we reverse the axes to improve the perspective. The algorithm began at point *A*, where the *β*^(0)^, *α*^(0)^, *V*^(0)^ = (0.4, 0.8, 3907). On iteration *k* = 10 at point *B*, where *β*^(10)^, *α*^(10)^, *V*^(10)^ = (0.446, 0.730, 101), the path of the algorithm had entered the ravine described in Figure 5 above. The path continued along the ravine until the stopping point at *β*^(40)^, *α*^(40)^, *V*^(40)^ = (0.233, 0.941, 76.7).

**Figure B1.**
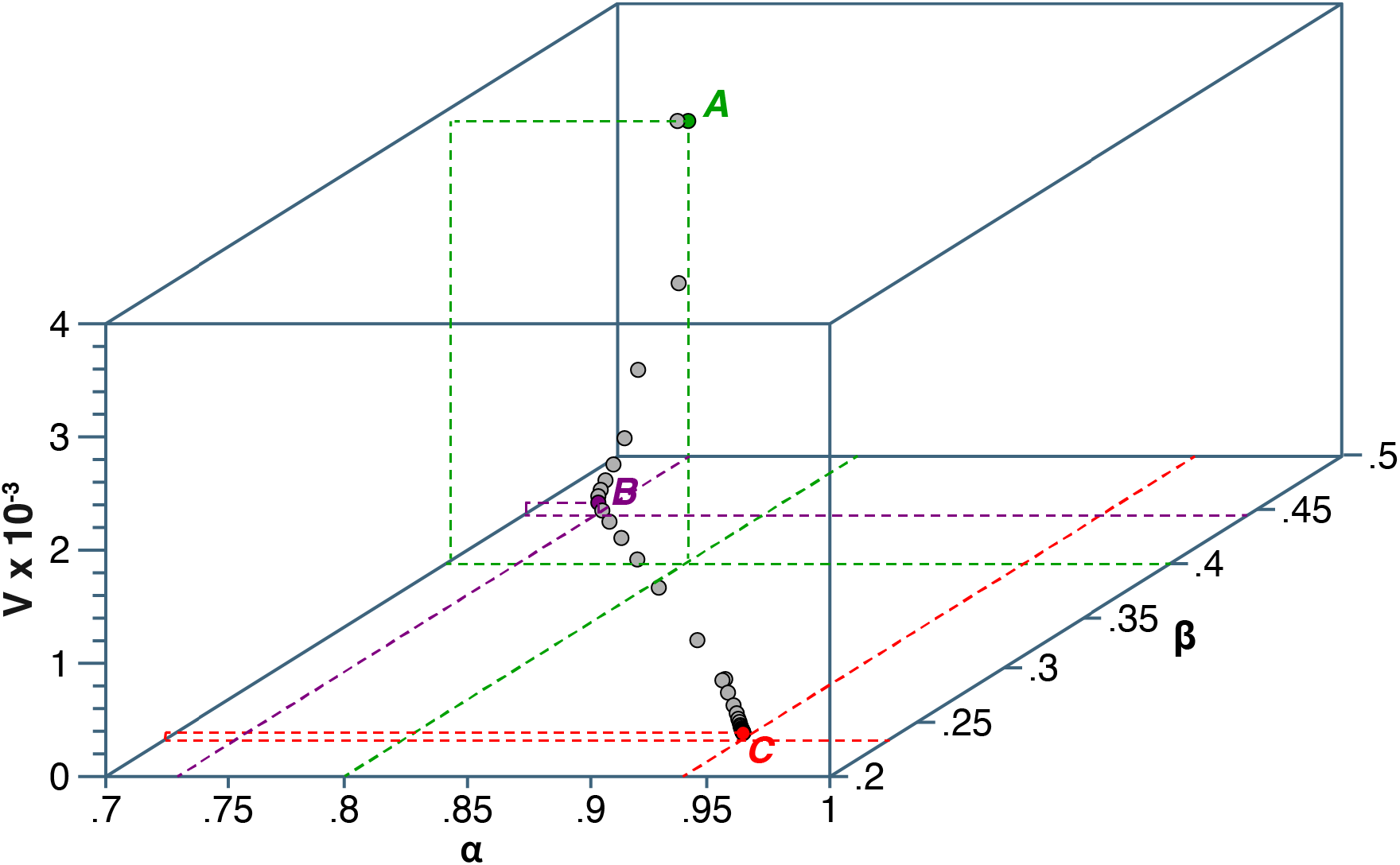
Path of (*β*^(*k*)^, *V*^(*k*)^, *V*^(*k*)^) Through Successive Iterations of the 4-Dimensional Newton-Raphson Algorithm: New York City Omicron Wave. The algorithm began at point *A*, where the *β*^(0)^, *α*^(0)^, *V*^(0)^ = (0.4, 0.8, 3907). The step size for successive iterations, defined in equation (16), was *q* = 0.5. At point *B*, where *β*^(10)^, *α*^(10)^, *V*^(10)^ = (0.446, 0.730, 101), the path of the algorithm had entered the ravine described in Figure 6. The path continued along the ravine until the stopping point at *β*^(40)^, *α*^(40)^, *V*^(40)^ = (0.233, 0.941, 76.7). The stopping criterion was | *V*^(*k*)^ − *V*^(*k*−1)^ | < 10^−4^. The execution time was 0.436 seconds.

## Declarations

### Sole Authorship

JEH is the sole author of this original work. He alone is responsible for the conceptualization of the work, the analysis of the data, the drafting of the manuscript, and the construction of the graphics. No requests for permission to use copyrighted material are required.

## Acknowledgments

The opinions expressed here are solely those of the author and do not necessarily reflect those of the Massachusetts Institute of Technology, Eisner Health, or any other organization or individual. I gratefully acknowledge the helpful comments of the following individuals on an earlier version of this manuscript: Julio R. Banga (Consejo Superior de Investigaciones Científicas, Spain), Keith Godfrey (University of Warwick, UK), Weiguo Han (University Corporation for Atmospheric Research, USA), Şiran Keske (Koç University School of Medicine, Turkey), Eduardo Massad (University of Sao Paolo, Brazil), Thomas Michelitsch (Sorbonne Université, France), Dimiter Prodanov (IMEC, Belgium), Emerson Sadurni (Benemérita Universidad Autónoma de Puebla, Mexico), Reinhard Schlickeiser (Ruhr-Universität Bochum, Germany), Boris Schmid (University of Oslo, Norway), and James A. Yorke (University of Maryland, USA). The comments of three anonymous referees are likewise appreciated.

## Earlier Versions

Earlier versions of this manuscript have been publicly posted on a preprint server [96].

## Human Subjects Declaration

This study relies exclusively on publicly available data that contain no individual identifiers. Links to the data sources are provided in the manuscript references.

## Competing Interests Declaration

The author has no competing interests.

## Funding Statement

This study did not receive any funding.

## Notes

### Competing Interest Statement

The authors have declared no competing interest.

### Summary of Updates

Additional explanation of the convergence properties of the proposed estimation method. Additional explanation of the application of the method to aggregate data on case incidence. Minor corrections to equations and references.

